# Structural damage of the cervical spinal cord within and beyond the maximal compression area in mucopolysaccharidosis types I and VI

**DOI:** 10.64898/2026.10.21.26365852

**Authors:** Igor Nestrašil, Christophe Lenglet, Ivan Krasovec, Bryon A Mueller, Alia Ahmed, Monica T Bondy, Ivana Labounková, Thierry Chekouo, Alena Svátková, Chester B Whitley, René Labounek

**Affiliations:** Division of Clinical Behavioral Neuroscience, Department of Pediatrics, University of Minnesota, Minneapolis, MN, USA; Center for Magnetic Resonance Research, Department of Radiology, University of Minnesota, Minneapolis, MN, USA; Kavli Institute for Systems Neuroscience, Trondheim, Trøndelag, Norway; Department of Psychiatry & Behavioral Sciences, University of Minnesota, Minneapolis, MN, USA; Division of Biostatistics & Health Data Science, School of Public Health, University of Minnesota, MN USA; Centre for Functional and Diagnostic Imaging, Department of Radiology, and DRCMR, Hvidovre and Amager Hospital, Denmark; Division of Genetics and Metabolism, Department of Pediatrics, University of Minnesota, Minneapolis, MN, USA

**Keywords:** . Mucopolysaccharidosis type I, Mucopolysaccharidosis type VI, Cervical myelopathy, Spinal cord compression, Magnetic resonance imaging (MRI), Diffusion tensor imaging (DTI), Neuroimaging biomarkers

## Abstract

**Background:** Cervical spinal cord (CSC) compression and myelopathy are frequent, severe complications in mucopolysaccharidosis (MPS) types I and VI. Early detection is critical to prevent irreversible neurological deficits and preserve quality of life. This prospective study aimed to identify quantitative neuroimaging biomarkers of structural CSC damage and myelopathy in MPS I and VI.

**Methods:** Structural and diffusion magnetic resonance imaging (MRI) of the CSC was analyzed in 17 MPS I patients, 3 MPS VI patients (19±7 years), and 19 case-matched healthy controls (20±7 years). CSC macrostructure (cross-sectional area, diameters) and microstructure (fractional anisotropy [FA], radial [RD], mean, axial diffusivities) were evaluated within and beyond the maximal compression areas.

**Results:** MPS patients demonstrated reduced CSC cross-sectional area and anterior-posterior diameter at upper cervical levels, alongside widespread microstructural alterations (lower FA, higher RD) throughout the CSC versus controls (p_FDR_<0.05). Restricted foramen magnum space was associated with microstructural damage at lower cervical levels without overt macrostructural compression. Morphological and microstructural metrics were sensitive to CSC compression and myelopathy severity (p_FDR_<0.05). Lower CSC integrity correlated with more advanced growth retardation (p<0.001). Among patients receiving hematopoietic stem cell transplantation, earlier treatment correlated with preserved upper CSC morphology (p<0.05). Distinct neuroimaging phenotypes were observed across MPS genotypes (p_FDR_<0.05).

**Conclusions:** CSC MRI provides candidate neuroimaging biomarkers for early detection and characterization of myelopathy in MPS I and VI. Structural damage extends beyond primary maximal compression areas, highlighting these markers for clinical monitoring and future therapeutic trials.

## Introduction

Mucopolysaccharidosis (MPS) disorders are a group of rare lysosomal storage disorders, each characterized by the deficiency of a specific lysosomal enzyme involved in glycosaminoglycan (GAG) degradation (Muenzer 2011; Nestrasil and Vedolin 2017). The resulting progressive accumulation of GAGs in cells and tissues leads to multi-organ dysfunction. All MPS disorders are inherited in an autosomal recessive manner, except MPS II, which is X-linked. Frequent manifestations of MPS include short stature, musculoskeletal abnormalities, hepatosplenomegaly, cardiorespiratory disease (Neufeld and Muenzer 2001; Muenzer 2011), and/or central nervous system (CNS) abnormalities (Neufeld and Muenzer 2001; Zafeiriou and Batzios 2013; Nestrasil and Vedolin 2017; Shapiro et al. 2017).

Cervical spine disorders are common in MPS and can cause cervical stenosis, leading to cervical spinal cord (CSC) compression, which may progress to CSC damage, resulting in irreversible neurological disability and severely affecting quality of life (Peck et al. 2016). MPS patients are at high risk of developing clinical myelopathy due to the high prevalence of cervical stenosis and CSC compression (Pantel et al. 2022; Costanzo et al. 2022; Kim et al. 2024). Cervical stenosis or CSC compression is common in MPS I (64-100%) (Stoop et al. 2013; Liu et al. 2022), IV (30-56%) (Solanki et al. 2013a, b; Akyol et al. 2019; Šáhó et al. 2026; Peene and Depreitere 2026), VI (80-100%) (Solanki et al. 2016; Kim et al. 2024), and VII (30-60%), and less common in MPS II (20-50%) (Scarpa et al. 2011; Pantel et al. 2022) and extremely rare in MPS III (White et al. 2011; Zafeiriou and Batzios 2013; Costanzo et al. 2022). In general, maximal compression usually occurs at the cranio-cervical junction or the first two cervical levels, i.e., C0-C2 (Costanzo et al. 2022).

MPS patients who develop myelopathy (Wald and Schmidek 1984; Clarke 1993; Costanzo et al. 2022) clinically manifest irreversible neurological symptoms and deficits, including pain and numbness in the limbs, poor coordination, imbalance, gait problems, and severe bladder problems, including incontinence (Wald and Schmidek 1984; Clarke 1993; Solanki et al. 2016; Davies et al. 2018; Costanzo et al. 2022). If untreated, myelopathy may lead to progressive disability, paralysis, quadriplegia, and, in the worst scenario, death (Crandall and Batzdorf 1966; Tetreault et al. 2017). Thus, early detection and timely intervention are imperative. Surgical treatment within the first 6 months improves clinical outcomes in degenerative cervical myelopathy (DCM) (Tetreault et al. 2015; Davies et al. 2017; Munro et al. 2023). However, only 20% of DCM cases are diagnosed early (Munro et al. 2023). Because clinical practice already favors prompt intervention, with 73% of MPS surgical treatments planned immediately upon myelopathy diagnosis (Costanzo et al. 2022), delivering surgical management within the critical 6-month window hinges entirely on early detection. Thus, establishing an early myelopathy diagnosis is vital to enable timely treatment and preserve patients’ long-term neurological function and quality of life.

Quantitative magnetic resonance imaging (MRI) of CSC morphology and microstructure identified candidates for neuroimaging markers sensitive to the early, presymptomatic stage of myelopathy and/or its rate of progression in DCM (Martin et al. 2017, 2018; Kadanka et al. 2017; David et al. 2019; Labounek et al. 2020; Valošek et al. 2021), traumatic spinal cord (SC) injury (David et al. 2019), adrenoleukodystrophy (Castellano et al. 2016; Huffnagel et al. 2019; van de Stadt et al. 2020), amyotrophic lateral sclerosis (Pisharady et al. 2020) and Friedreich’s ataxia (Joers et al. 2022; Rezende et al. 2023). To further advance early myelopathy detection, the current prospective study aimed to identify candidates for myelopathy neuroimaging biomarkers (Strimbu and Tavel 2010; Feng and Pepe 2020) in MPS I and VI and tested 3 working hypotheses:

*Hypothesis 1: CSC structure is damaged beyond the C0-C2 segment in MPS I and VI*.

*Hypothesis 2: CSC structure is sensitive to the severity of compression and myelopathy in MPS I and VI*.

*Hypothesis 3: Quantitative neuroimaging of CSC structure provides imaging phenotypes that are sensitive to distinct MPS genotypes*.

## Materials and Methods

### Study Design and Recruitment

This study prospectively collected multi-parametric quantitative MRI data of the CSC from a subset of participants enrolled in the Lysosomal Disease Network (LDN; https://ldn.rarediseasesnetwork.org), a collaborative clinical research consortium dedicated to investigating rare, inherited lysosomal storage disorders.

### Participant Recruitment and Ethical Considerations

The dataset was derived from a subset of patients and volunteers recruited within the broader LDN registry framework between August 1, 2016, and December 31, 2024. Prior to the initial in vivo MR examinations, the study protocols were approved by the University of Minnesota Institutional Review Board (STUDY00014153, STUDY00004912, and STUDY00022289). Written informed consent was obtained from all subjects or their legal guardians before participation

### Clinical and Surgical Data Collection

We extracted clinical, demographic, and surgical data from the institutional REDCap database for all participants. Recorded variables included age, sex, body mass index (BMI), and specific MPS subtype (MPS I-H [Hurler], I-HS [Hurler-Scheie], I-S [Scheie], and MPS VI [Maroteaux-Lamy]). Overall somatic disease burden was quantified at each clinical visit using the Mucopolysaccharidosis (MPS)-specific Physical Symptom Score (PSS) (Ahmed et al. 2016a, b).

Medical histories were reviewed to document disease-modifying treatments and surgical interventions. For patients treated with hematopoietic stem cell transplantation (HSCT), we recorded transplant modality (bone marrow [BMT] or cord blood [CBT]), donor match status, and age at transplantation. Detailed neurosurgical and orthopedic histories captured the type, anatomical levels, and patient age for all prior spinal surgeries (e.g., cervical or thoracolumbar fusion, laminectomy, decompression, and osteotomy).

### Neurological and Orthopedic Evaluation

Clinical neurological and musculoskeletal status at the scan date was systematically documented. Orthopedic evaluations recorded short stature, scoliosis, and the specific level of kyphosis (cervical, thoracic, or lumbar). Neurological assessments evaluated abnormal reflexes, carpal tunnel syndrome, and hydrocephalus (distinguishing active hydrocephalus from passive hydrocephalus *ex vacuo*). Gait abnormalities were differentiated as neurologically driven or secondary to orthopedic deformities. Specific anatomical anomalies, including ligament hypertrophy and odontoid capping, were evaluated through a consensus reading integrating medical history with MRI findings.

### Mutation Analysis

We retrieved genetic diagnostic data from individual medical records to characterize each participant’s genotype. Documented genetic variables included the specific primary and secondary allelic mutations (e.g., W402X, Q70X, L238Q). Mutations were further categorized by zygosity (homozygous or heterozygous) and variant consequence, distinguishing among nonsense, missense, deletion, and splice-site mutations to accurately determine the genotype-phenotype severity.

### MRI Acquisition

Sagittal T_2_-weighted (TR/TE 3500/91ms, voxel size 0.7×0.7×3.3mm^3^), T_1_-weighted (TR/TE 4000/90ms, voxel size 0.7×0.7×3.3mm^3^) and axial T_2_-weighted (TR/TE 4000/90ms, voxel size 0.56×0.56×3.3mm^3^; 30-36 slices) scans followed by diffusion MRI (dMRI) acquisition were acquired on a 3T Siemens Prisma MR scanner, at the Center for Magnetic Resonance Research (CMRR), University of Minnesota, using a 20-channel head/neck coil. The field of view (FOV) of the axial T2-weighted scans was positioned to cover cervical levels C1–C7, with the SC centered in the slice. In the sagittal plane, the FOV was oriented as perpendicular to the SC as possible. To match the FOV centering and orientation of the preceding axial anatomical scan, dMRI was performed using a RESOLVE (REadout Segmentation Of Long Variable Echo trains) sequence with two identical runs in opposite phase-encoding directions (anterior–posterior and posterior–anterior). Thirty diffusion-weighted volumes (b=650smm^-2^) and 6 b_0_ images covering levels C1–C7 were acquired across 30–36 contiguous axial slices (TR=4500ms, TE_1_ =52ms, TE_2_ =82ms, voxel size 1.1×1.1×3.3mm^3^).

### Radiological Reading

An experienced radiologist (A.S.) evaluated all structural MRI scans. The core protocol reviewed included sagittal T_1_-weighted turbo spin-echo (TSE), sagittal T_2_-weighted TSE, sagittal T_2_-weighted fluid-sensitive sequences (TIRM or STIR), and axial T2-weighted TSE scans. When available, axial T_2_*-gradient-echo (MEDIC) and motion-insensitive radial sequences (T2 BLADE) were used to mitigate motion or hardware artifacts. The diagnostic field of view extended from the skull base down to the upper/mid-thoracic spine (T4–T7). Longitudinal scans across available time points were systematically evaluated for:

● **Stenosis and Cord Compression:** Spinal canal stenosis, ventral cerebrospinal fluid (CSF) space narrowing, anatomical levels of maximal cord compression, and contributing factors (disc protrusion/prolapse, ligamentous hypertrophy).
● **Myelopathy:** Pathological SC T2/TIRM/STIR hyperintensities (e.g., "owl-eyes" sign), carefully distinguishing true myelopathic signal from motion, swallowing, or metallic susceptibility artifacts from prior stabilization surgeries (osteosynthesis).
● **Additional Pathologies:** Secondary findings, including syringomyelia, vertebral hemangiomas, and post-surgical structural changes.

### Image Analysis

Sagittal and axial T_2_-weighted anatomical scans were bias-field corrected (Tustison et al. 2010). The sagittal T_2_-weighted scan was resampled to an isotropic voxel size, and the SC was segmented using the `deepseg_sc` algorithm.(Gros et al. 2019) The locations of intervertebral discs were manually labeled in the resampled sagittal T_2_-weighted scan using `sct_label_utils` implemented in the Spinal Cord Toolbox (version 6.1) (De Leener et al. 2017). The SC was also segmented (Gros et al. 2019) in the T_2_-weighted axial anatomical scan. Sagittal and axial T_2_-weighted anatomical scans were co-registered (De Leener et al. 2018), and the locations of cervical levels were warped into the space of the T_2_-weighted axial scan. Level-specific morphology of the CSC was assessed with `sct_process_segmentation` using angle correction as implemented in the SCT (De Leener et al. 2017).

Susceptibility, motion, and eddy current artifacts were suppressed in dMRI data using topup(Andersson et al. 2003) and eddy(Andersson and Sotiropoulos 2016) algorithms implemented in the FMRIB Software Library (FSL version 6.0.1) (Jenkinson et al. 2012). A diffusion tensor (Basser et al. 1994) was fitted in each image voxel. An averaged diffusion-weighted image (DWI) and a b_0_ scan were estimated from the preprocessed dMRI data. The SC was segmented (Gros et al. 2019) from the averaged DWI. Using the dMRI data SC mask, the averaged b_0_ scan was co-registered (De Leener et al. 2018) with the axial T_2_-weighted anatomical scan and cervical level labels were warped into the dMRI data space. Level-specific FA, MD, AD, and RD measurements were estimated as the weighted average across the entire SC cross-section (Lévy et al. 2015).

### Occipito-cervical Junction Measurements

Measurements of occipito-cervical junction morphology were obtained using the ITK-Snap (Yushkevich et al. 2006) biomedical imaging software suite (www.itksnap.org). All measurements, except for the cord diameter (CD) and foramen magnum diameter (FM) were obtained from the participant’s midsagittal T_1_-weighted image. CD and FM were measured on the axial plane of the T_2_-weighted image below the foramen magnum. Measurements of Power’s Ratio (PR), atlantodental interval (ADI), basion-dens interval (BDI), basion-axial interval (BAI), and clivus-canal angle (CCA) were obtained following the methods of (Rojas et al. 2007). The dens-to-laminar interval (D2L) was measured as the distance from the most posterior aspect of the dens to the lamina of the spinal canal. The rater providing the manual measurements (IK) was blinded to the clinical status of the analyzed images.

### Statistical Analysis

Two-sample t-tests assessed between-group differences in structural MRI measurements (p_FDR_<0.05; corrected for the false discovery rate using adjusted Benjamini-Hochberg p-values). Post-hoc analysis of covariance (ANCOVA; p<0.05) with body weight or body height (Labounek et al. 2025) as a confounding factor was performed for structural MRI measurements where the preceding t-test identified significant differences. Cross-correlation analysis using the Pearson correlation coefficient identified strongly intra-correlated clusters in structural MRI measurements. Strongly intra-correlated clusters were substituted with cluster-specific averages to reduce data dimensionality. A Pearson correlation coefficient matrix was estimated for the reduced dataset, and variables were rotated into the orthogonal space of the principal components. Hidden inter-variable relationships were identified in the rotated space. In MPS I patients with Hurler syndrome and MPS VI patients, neuroimaging measurements correlated with age at HSCT (p<0.05); ANCOVA adjusting for age at MRI scan acquisition as a confounder further confirmed the relationship (p<0.05). Two-sample t-tests (p_FDR_<0.05) further investigated the sensitivity of neuroimaging markers to MPS genotypes and to the severity of SC compression and myelopathy. Frequencies of other pathological findings in our MPS cohort were also assessed.

## Results

### Participant characteristics

Longitudinal high-resolution structural MRI and dMRI covering the entire CSC (i.e., C1-C7 levels) were acquired in 17 MPS I patients, 3 MPS VI patients (9 females, 11 males; body height 133±16 cm; body weight 43±20 kg), and 19 case-matched (i.e., sex-, age-, and BMI-matched) healthy controls (HC; 8 females, 11 males; body height 164±16 cm; body weight 61±19 kg; cross-sectional data only). Baseline matching demographics are provided in **Table 1**. The study cohort differed significantly between groups in body height (p<0.001) and body weight (p=0.005). The Physical Symptom Score (PSS) in the recruited cohort of MPS patients ranged from 4 to 14, with a median value of 10 (**Table 1**). Mutation analysis in 17 MPS I patients identified 13 homozygotes with Hurler syndrome, 3 heterozygotes with Hurler-Scheie syndrome, and 1 heterozygote with Scheie syndrome (**Table 1**). Thirteen patients with Hurler syndrome and 2 MPS VI patients were treated with HSCT at the age of 16±8 months. Seven patients received cord blood stem cells (14±7 months old; donor: unrelated match in 5 cases, sibling in 1 case, and unknown in 1 case) and another 8 received bone marrow stem cells (18±8 months old; donor: unrelated match in 3 cases, sibling in 5 cases; **Table 1**).

**Table 1:**
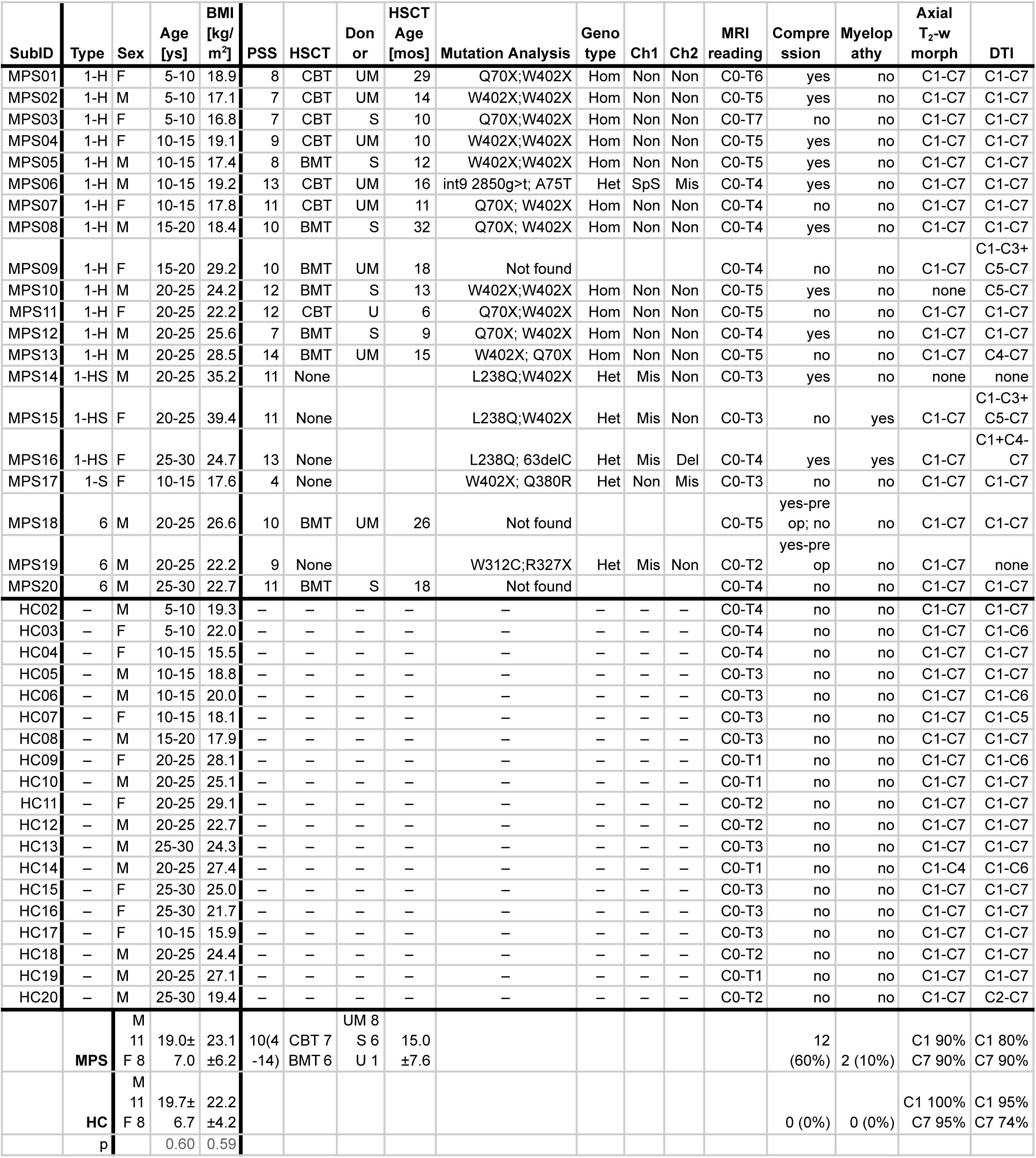
Study cohort demographics and cervical spinal cord imaging availability. MPS02-HC02; MPS03-HC03; etc. are the case-matched pairs. *<u>Abbreviations:</u>* SubID - subject identifier; MPS - mucopolysaccharidosis; HC - healthy control; M - male; F - female; ys - years; BMI - body mass index; PSS - physical symptom score; MRI - magnetic resonance imaging; DTI - diffusion tensor imaging; p - p-value of two-sample t-test; C1-C7 - range of cervical levels 1-7; H - Hurler syndrome; HS - Hurler-Scheie syndrome; S - Scheie syndrome; HCST - Hematopoietic Stem Cell Transplantation; mos - months; BMT - Bone Marrow Transplant; CBT - Cord Blood Transplant; UM - Unrelated Match; S - Sibling; U - Unknown; Het - Heterozygous; Hom - Homozygous; Ch1 - Chromosome 1 mutation; Ch2 - Chromosome 2 mutation; SpS - Splice site; Mis - Missense; Non - Nonsense. The group-specific averages are presented as mean ± standard deviation or as median(minimum-maximum).

All MPS patients had stenosis, with the most frequent occurrences at the C0-C2 levels. However, stenosis in the thoracic and lumbar segments was also quite common. Degenerative SC compression was present in 12 (60%) of enrolled MPS cases. Eight (62%) had compression in Hurler syndrome; two (67%) in Hurler-Scheie syndrome; and two (67%) were surgically treated in MPS VI. Myelopathy was present in 2 (10%) of enrolled cases. Both myelopathic cases belonged to the Hurler-Scheie syndrome, suggesting a myelopathy prevalence of up to 67% in this MPS I genotype. Subject MPS15 (**Table 1**) demonstrated clinical myelopathy that worsened after a fall. Subject MPS16 (**Table 1**) demonstrated radiological and clinical myelopathy with no history of severe trauma.

Twelve (60%) MPS patients had a history of SC surgery. Four (31%) patients with Hurler syndrome underwent surgery 2-7 years after the MRI in this study and 3 (23%) underwent it 11-22 years before the study MRI. Three (100%) patients with Hurler-Scheie syndrome underwent surgery (one year after the study MRI and two patients 6-7 years before the study MRI). Moreover, the patient who underwent surgery 6 years before the MRI had a second SC surgery 3 years after the MRI. Two (67%) patients with MPS VI had surgery 1 and 7 years before the MRI, respectively. **Table 2** provides more details on the spinal surgeries.

**Table 2:**
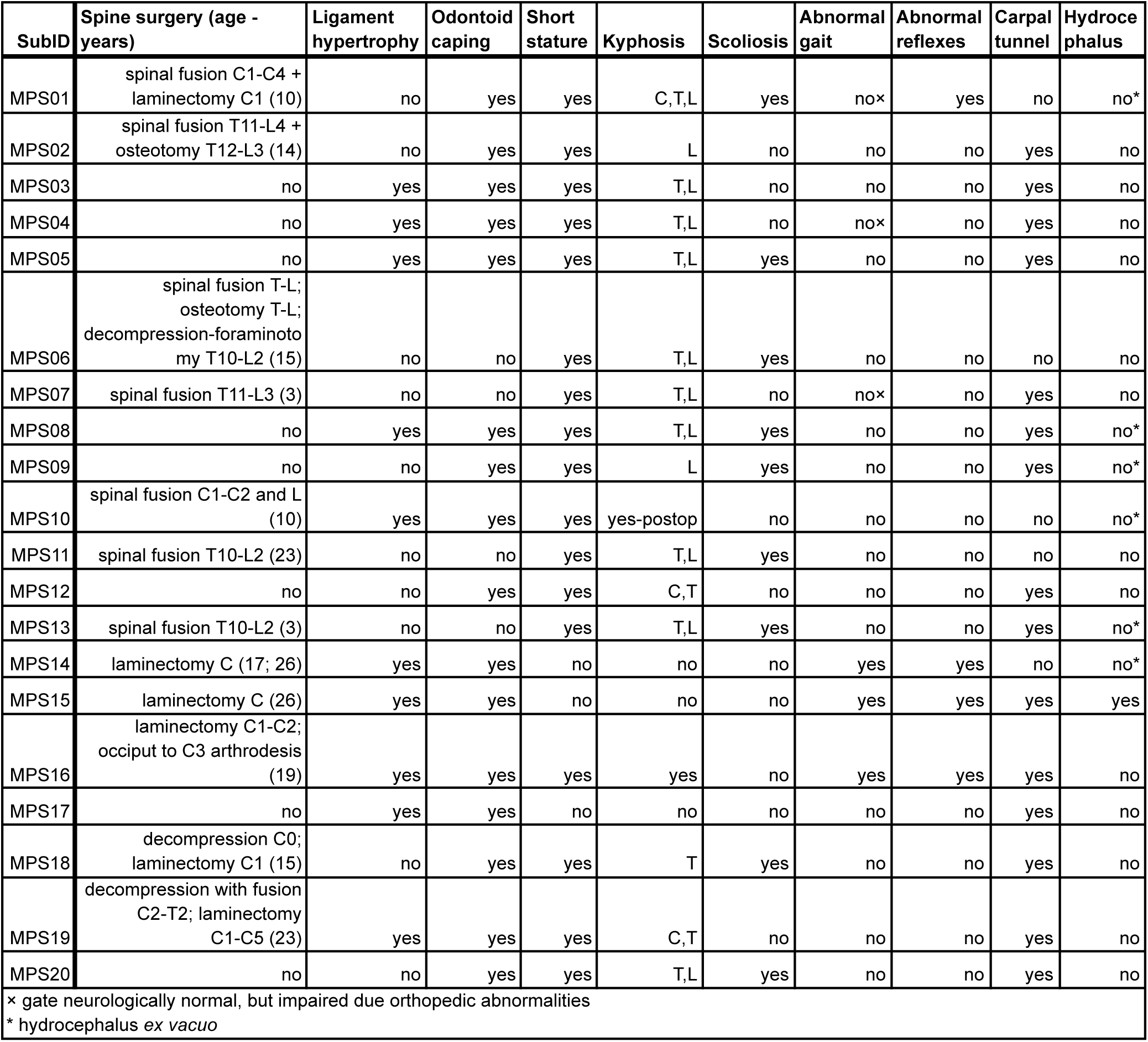
Clinical records in enrolled MPS patients.

Ten (50%) patients had ligament hypertrophy; 16 (80%) had odontoid capping; 17 (85%) had short stature (i.e., height percentile <3%); 17 (85%) had gibbus or kyphosis; 9 (45%) had scoliosis; 3 (15%) demonstrated abnormal gait; 3 (15%) had impaired gait due to orthopedic abnormalities; 4 (20%) had abnormal reflexes; 15 (75%) were treated for carpal tunnel; 1 (5%) had hydrocephalus and an additional 6 (30%) had hydrocephalus *ex vacuo*. Patient-specific clinical records are summarized in **Table 2**.

### Image quality for quantitative MRI

Representative examples of sagittal and axial T_2_-weighted anatomical scans and DTI-based (diffusion tensor imaging) maps of CSC microstructure are shown for a case-matched healthy control (**Fig. 1a**) and an MPS I patient (**Fig. 1b**). Cervical vertebrae and intervertebral disc deformity along with characteristic narrowing at the occipito-cervical junction are visible in the sagittal anatomical scan (**Fig. 1b**). All DTI-based diffusivity maps (i.e., axial diffusivity - AD; mean diffusivity - MD; and radial diffusivity - RD) showed clear contrast between SC tissue and cerebrospinal fluid (**Fig. 1a,b**). Moreover, all diffusivity maps showed image contrast in other structures, such as nerve roots and vertebrae, matching the shapes and locations in the corresponding high-resolution axial anatomical scans (**Fig. 1a,b**). The fractional anisotropy (FA) map demonstrated expected domain values of 0<FA<1 in SC tissue, with darker (lower) FA values in areas of expected gray matter (**Fig. 1a,b**). Automated CSC morphology was assessed using axial T_2_-weighted anatomical scans (**Fig. 1a,b**). Manual measurements of morphological integrity at the occipito-cervical junction used sagittal T_1_-weighted and T_2_-weighted anatomical scans and axial T_2_-weighted anatomical scans (**Fig. 1a,b**; **Fig. 2a**).

**Figure 1:**
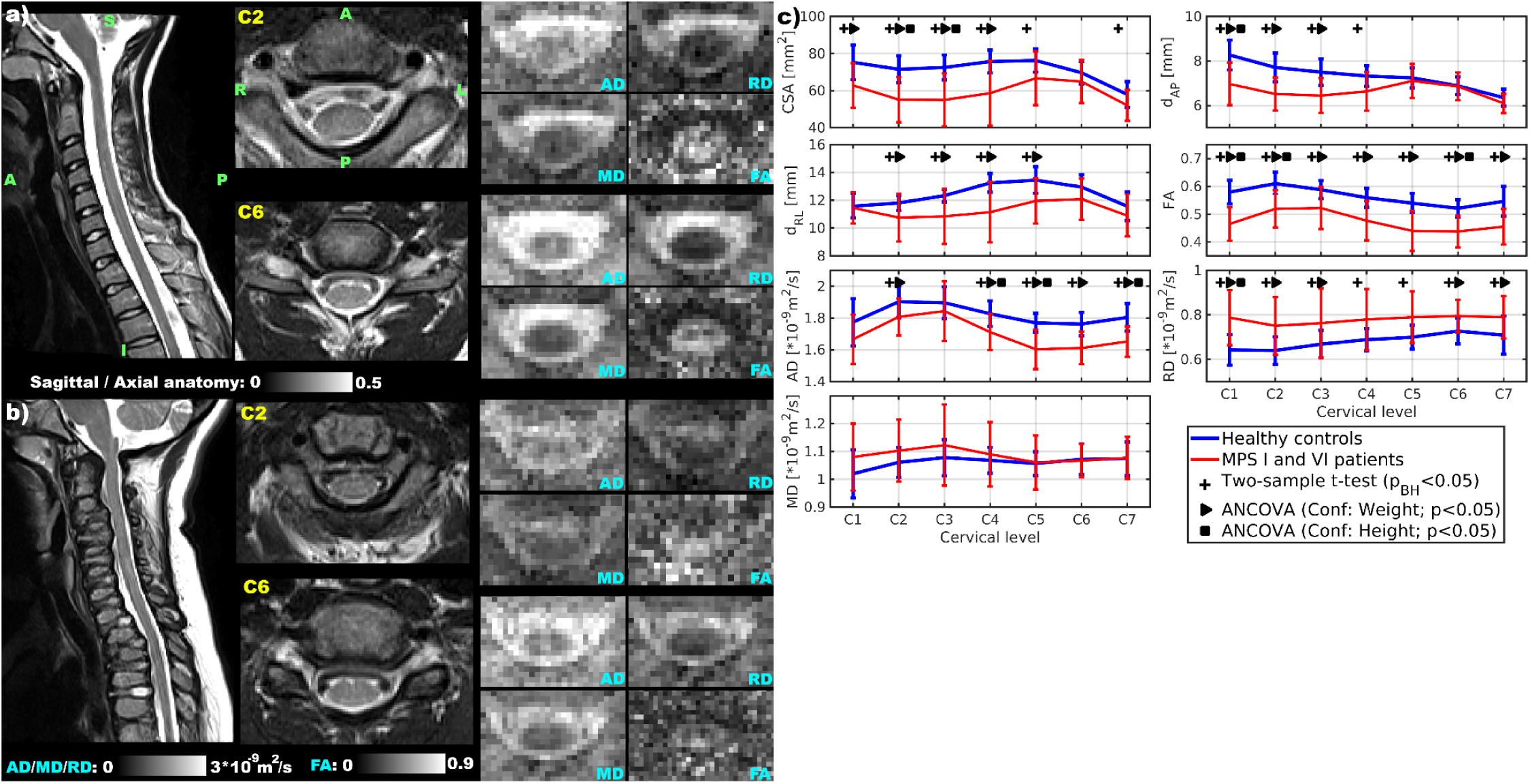
Altered cervical spinal cord morphology and microstructure in MPS I and VI. **a**, contributes to microstructural myelopathy in SC areas distant from the cervical maximal compression levels T_2_-weighted anatomical scan. The orientation of all shown sagittal slices is as denoted: S - superior; I - inferior; A - anterior; and P - posterior. In the middle are two representative axial slices from the high-resolution T_2_-weighted anatomical scan at the cervical C2 and C6 levels. The orientation of all shown axial slices is as denoted for the C2 axial anatomical scan: A, P, L - left; and R - right. On the right are microstructural maps derived with diffusion tensor imaging (DTI) for the same C2 and C6 slices as shown in the middle with the axial T_2_-weighted anatomical scan. The microstructural maps are fractional anisotropy (FA), axial (AD), mean (MD), and radial (RD) diffusivity. **b**, Representative imaging example of a female MPS I patient. The shown images and their order are identical to those shown in panel **a**. **c**, Error bars of morphological and microstructural measurements in cervical levels C1-C7 for groups of case-matched healthy controls and MPS I and VI patients. Each error bar shows a group-specific mean and a ± one standard deviation interval. Cross markers above error bars indicate measurements in which two-sample t-tests with adjusted Benjamini-Hochberg p-values (*p_BH_<0.05*) identified significant between-group differences. Triangle and square markers above error bars indicate measurements in which post-hoc analyses of covariance (ANCOVAs) using body weight or body height as the confounding factor identified significant between-group differences (p<0.05). The spinal cord morphological measurements were cross-sectional area (CSA), anterior-posterior diameter (d_AP_) and right-left diameter (d_RL_).

**Figure 2:**
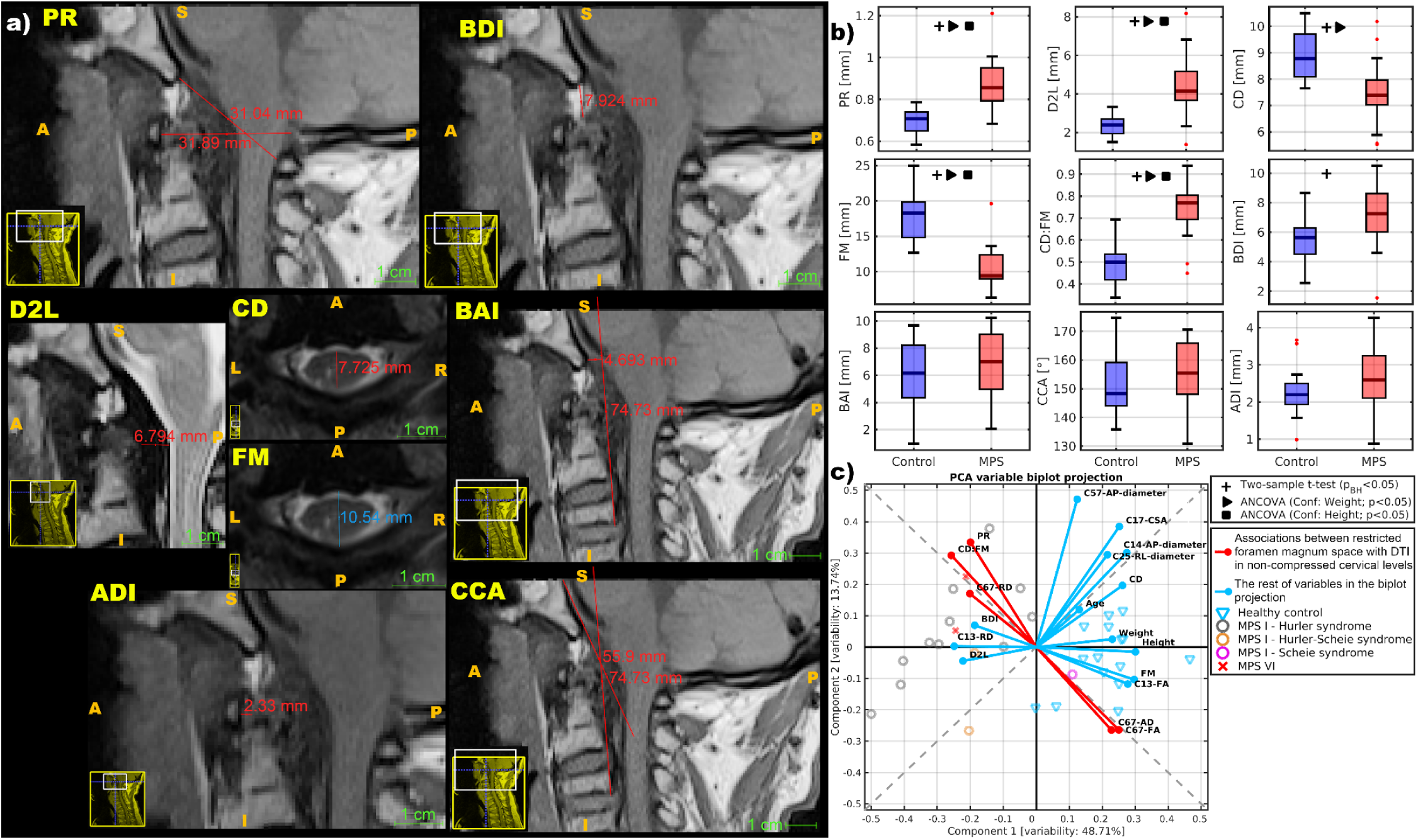
Altered structural integrity of the occipito-cervical junction and associations between restricted foramen magnum space and loss of inner integrity at non-compressed low cervical levels. **a**, Representative example of manual measurements shown in T_1_-weighted sagittal and T_2_-weighted axial scans for a male MPS VI patient. The manual measurements were Power’s Ratio (PR), basion-dens interval (BDI), dens-to-laminar interval (D2L), cord diameter (CD), foramen magnum diameter (FM), basion-axial interval (BAI), atlantodental interval (ADI), and clivus-canal angle (CCA). CD and FM were measured just below the foramen magnum. Orientation of each image is as denoted: S - superior; I - inferior; A - anterior; P - posterior; L - left; R - right. **b**, Group-specific boxplots of the manual measurements in the occipito-cervical junction area. The MPS group included both MPS I and VI patients. Cross markers above the boxplots indicate measurements in which two-sample t-tests using adjusted Benjamini-Hochberg p-values (*p_BH_<0.05*) identified significant differences between groups. Triangle and square markers above the boxplots indicate measurements in which post hoc analyses of covariance (ANCOVAs) using body weight or body height as the confounding factor identified significant differences between groups (p<0.05). **c**, The biplot projection of the 1^st^ and 2^nd^ principal components, applied to the reduced variable space, shows (red-color highlighted) associations between restricted foramen magnum space (i.e., higher CD:FM ratio) and higher radial diffusivity (C67-RD), lower fractional anisotropy (C67-FA) and lower axial diffusivity (C67-AD) in the non-compressed cervical levels C6-7. The biplot projection also shows that body height, body weight, and age have minimal effects in this association. Scores for participants with complete records are projected onto the first two principal components, demonstrating clear separation between healthy controls and MPS patients, except for the Scheie syndrome patient, who overlaps with controls.

Automated measurements of CSC morphology were unavailable for 2 MPS I male patients due to low-quality anatomical MR images (**Table 1**). One male HC lacked CSC morphology measurements for the C5-C7 segments (**Table 1**).

Automated measurements of CSC microstructure were unavailable for 1 MPS I and 1 MPS VI male patients due to low-quality dMRI images (**Table 1**). For two additional male MPS I patients, only C4-C7 and C5-C7 dMRI measurements were available (**Table 1**). Fat-induced artifacts in dMRI data were present at the C4 level for two female MPS I patients and at the C2-C3 level for one additional female MPS I patient (**Table 1**). Artifact-affected cervical levels were excluded from statistical analyses. One male HC had a missing dMRI measurement in the C1 segment; one female HC in the C6-C7 segment; and two male and two female HCs in the C7 segment (**Table 1**).

Longitudinal MRI scans were available for 5 MPS I and 1 MPS VI patients. One male MPS I patient had an additional data point acquired 1 year before the case-matched visit. One female and one male MPS I patient had an additional data point acquired 1 year after the case-matched visit. One female MPS I and one male MPS VI patient had additional data points acquired 1 year before and 1 year after the case-matched visit. Finally, one male MPS I patient had additional data points acquired 2 and 3 years after the case-matched visit. All longitudinal and subject-specific MRI measurements are shown in **Supplementary Figures S1-S8**.

### CSC morphology and microstructure are altered in MPS I and VI

The CSC cross-sectional area (CSA) was lower in the MPS I and VI patient group at the C1-C4 levels (**Fig. 1c**). The anterior-posterior diameter (d_AP_) was lower at the C1-C3 levels (**Fig. 1c**). The right-left diameter (d_RL_) was lower at the C2-C5 levels (**Fig. 1c**). FA was lower and RD higher across the entire CSC (**Fig. 1c**). AD was lower at the C4-C7 levels (**Fig. 1c**). All subject-specific measurements, including available longitudinal measurements, are shown in **Supplementary Figures S1-S7**. These supplementary plots demonstrate the degree of overlap between MPS I and VI measurements. However, genotype-specific nuances in these neuroimaging markers are reported further in the text.

### Structural integrity of the occipito-cervical junction area is altered in MPS I and VI

Representative examples of manual radiologic measurements are shown in **Fig. 2a**. The cord diameter (CD) and foramen magnum diameter (FM) were lower in MPS patients (**Fig. 2b**). The Power’s ratio (PR), dens-to-laminar interval (D2L), and CD:FM ratio were higher in MPS patients (**Fig. 2b**). The basion-dens interval demonstrated a trend toward higher values in MPS patients (**Fig. 2b**). There were no differences in basion-axial interval (BAI), clivus-canal angle (CCA) and atlantodental interval (ADI) between MPS patients and HC (**Fig. 2b**). All subject-specific measurements, including available MPS longitudinal measurements, are shown in the **Supplementary Figure S8**.

### Obstruction in the foramen magnum interacts with inner integrity disruption in the lower CSC

Quantitative imaging and a limited sample size yielded a large number of variables that formed highly intra-correlated clusters (**Supplementary Figure S9**). Therefore, we reduced the variable space to 18 variables, with some variables represented as averages of the identified highly intra-correlated clusters (**Supplementary Figure S9**; **Fig. 2c**). Rotation of the dataset into the space of the first two orthogonal principal axes revealed an interaction between restricted foramen magnum space (i.e., the CD:FM ratio) and DTI-based microstructural integrity disruption (i.e., C67-FA, C67-AD and C67-RD) at low CSC levels (**Fig. 2c**), where macrostructural (morphological) changes were not observed (**Fig. 1c**). Therefore, we present objective evidence that MPS-related compression at the occipito-cervical junction (**Fig. 2b**) contributes to microstructural myelopathy in SC areas distant from the cervical maximal compression levels (**Fig. 1c**; **Fig. 2c**). Moreover, the data rotation into the two principal axes showed clear separation between HC and MPS patients, except for the Scheie syndrome patient (**Fig. 2c**). Overall, the data objectively confirms the first hypothesis that CSC structure is damaged beyond the C0-C2 segment in MPS I and VI.

### Sensitivity of CSC neuroimaging markers to disease severity, therapy response, and patients’ genotype

CSA and d_AP_ were lower in MPS patients with stenosis than in HC (**Fig. 3a**). CSA and d_RL_ were lower in MPS patients with SC compression than in those with stenosis (**Fig. 3a**). CSA and d_RL_ were lower in MPS patients with myelopathy when compared to MPS patients with compression (**Fig. 3a**). Notably, the two patients with Hurler-Scheie syndrome and myelopathy were young adults. Yet, they exhibited the smallest CSA morphology in the recruited cohort, with ages 7-30 years. Effect sizes for the myelopathy severity comparisons are summarized in the first panel of **Table 3**. Among the manual craniocervical measurements, Power’s ratio, dens-to-laminar interval (D2L), foramen magnum diameter, cord-to-foramen magnum ratio (CD:FM), and basion-dens interval (BDI) effectively distinguished patients with stenosis from HC (**Table 3**).

**Figure 3:**
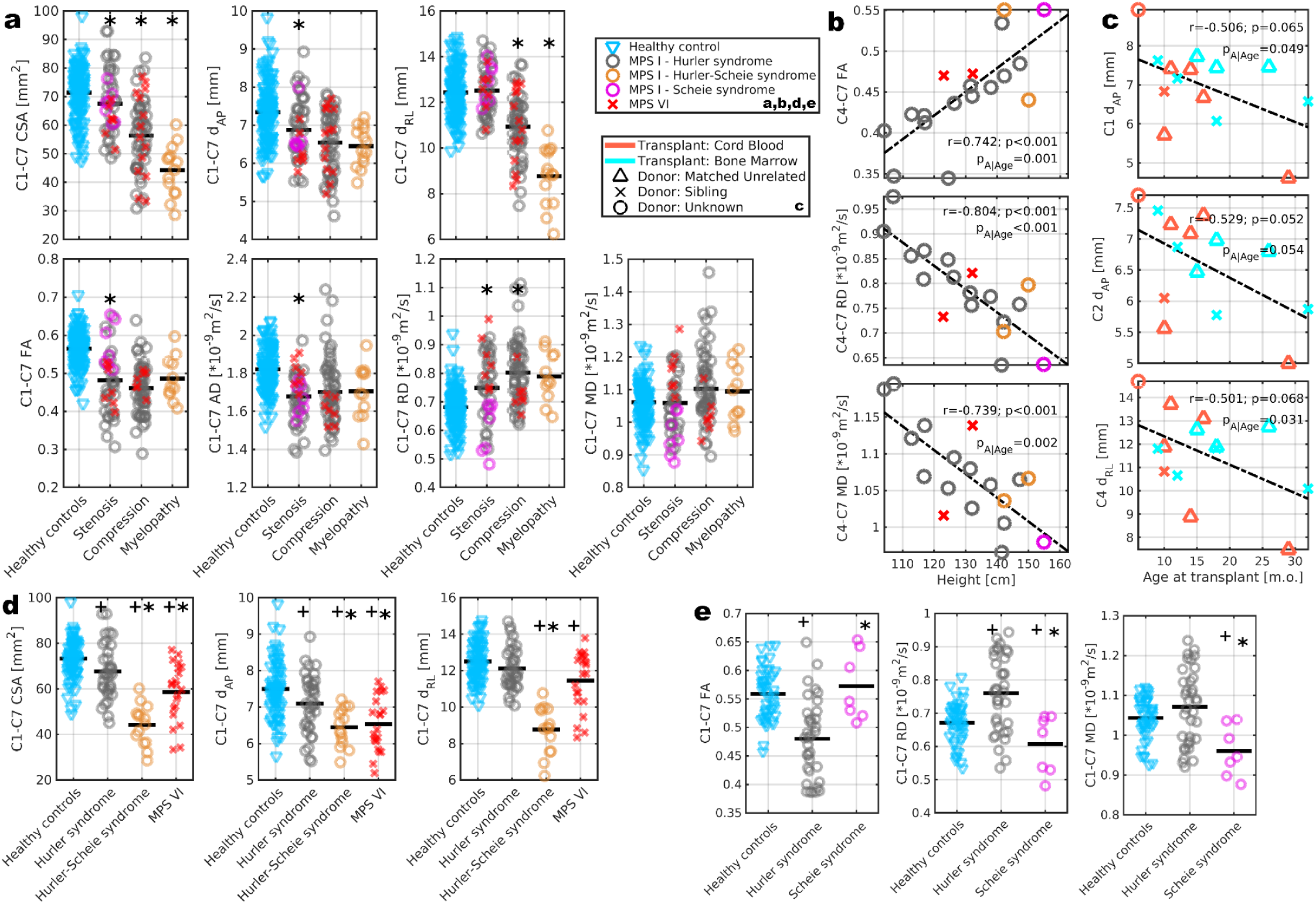
Cervical neuroimaging phenotypes are sensitive to the severity of spinal cord compression and myelopathy (a), growth deficits (b), treatment timing (c), and MPS genotypes (d,e) **a**, Sensitivity of cervical spinal cord (CSC) morphology and diffusion tensor imaging (DTI) to the severity of spinal cord (SC) compression and myelopathy. Investigated measurements were aggregated from C1 to C7 levels. Abbreviations of investigated measurements are: CSA, cross-sectional area; d_AP_ anterior-posterior diameter; d_RL_ right-left diameter; FA fractional anisotropy; AD axial diffusivity; RD radial diffusivity; and MD mean diffusivity. A star indicates measurements where two-sample t-tests identified significant between-group differences (p_FDR_<0.05). MPS patients with stenosis were compared to healthy controls (HC); MPS patients with compression were compared to patients with stenosis; and MPS patients with myelopathy were compared to patients with compression. **b**, Pearson correlation coefficients (*r*) with corresponding p-value (*p*) between mid and lower CSC DTI and body height in MPS I and VI patients. Measurements were averaged from C4 to C7 levels. Supplementary material shows that such correlations with body height were not observed in the HC. The *p_A|Age_*shows the p-value of the relationship when age at scan is used as a confounding factor in analysis of covariance (**b**, **c**). **c**, Causality between age at treatment (m.o. - months old) and upper CSC morphology. **d**,**e** Aggregated genotype-specific neuroimaging measurements. The categorical scatter plots show subsets of age-matched data samples (i.e., three 20-30-year-old patients with Hurler-Scheie syndrome; three 20-30-year-old MPS VI patients; and one patient with Scheie syndrome). The between-genotype differences were compared to Hurler syndrome data with two-sample t-tests (adjusted Benjamini-Hochberg p-values *p_BH_<0.05*) and marked with a star above the other genotypes that differed in morphological (**d**) and DTI measurements (**e**). The cross markers identify MPS genotypes whose measurements differed when compared to HC with two-sample t-tests (*p_BH_<0.05*; **d,e**). Investigated measurements were aggregated from C1 to C7 levels.

**Table 3.** Effect sizes (*Cohen’s d*) for cervical spinal cord quantitative MRI parameters across myelopathy severity categories and MPS genotypes Effect sizes (*Cohen’s d*) and corresponding 95% confidence intervals (95% CI, shown in parentheses) for spinal cord morphological and microstructural measurements. Comparisons are stratified into three analytical panels: 1. *Myelopathy severity classification across the full cohort:* Pairwise comparisons between healthy controls and patients grouped by disease severity (stenosis, degenerative spinal cord compression, and clinical myelopathy). 2. *Genotype-phenotype analysis in adult and late-adolescent participants (>16.5 years old):* Pairwise comparisons among controls, MPS I-H (Hurler), MPS I-HS (Hurler-Scheie), and MPS VI subcohorts. 3. *Genotype-phenotype analysis in pediatric/younger participants (7.5–15.5 years):* Pairwise comparisons among age-matched controls, MPS I-H (Hurler), and MPS I-S (Scheie) subcohorts. An asterisk (*) indicates statistically significant differences between groups based on a two-sample t-test with False Discovery Rate correction using the Benjamini–Hochberg procedure (*p_BH_<0.05*). *Abbreviations:* AP, anterior-posterior; RL, right-left; D2L, dens-to-laminar interval; CD:FM, ratio of cord diameter to foramen magnum diameter; BDI, basion-dens interval; MPS, mucopolysaccharidosis.

| **Myelopathy severity**<br>(all participants) | Controls vs.<br>Stenosis | Stenosis vs.<br>Compression | Compression vs.<br>Myelopathy |
| --- | --- | --- | --- |
| Cross-sectional area | 0.40 (0.07; 0.73)* | 0.91 (0.52; 1.30)* | 0.99 (0.39; 1.58)* |
| AP diameter | 0.60 (0.26; 0.93)* | 0.40 (0.03; 0.78) | 0.12 (-0.45; 0.69) |
| RL diameter | -0.08 (-0.41; 0.25) | 1.23 (0.83; 1.64)* | 1.53 (0.91; 2.15)* |
| Fractional anisotropy | 1.39 (1.02; 1.76)* | 0.27 (-0.12; 0.66) | -0.39 (-1.02; 0.25) |
| Axial diffusivity | 1.24 (0.87; 1.60)* | -0.16 (-0.54; 0.23) | -0.02 (-0.65; 0.62) |
| Radial diffusivity | -0.78 (-1.13; -0.43)* | -0.45 (-0.84; -0.06)* | 0.12 (-0.52; 0.76) |
| Mean diffusivity | 0.03 (-0.31; 0.37) | -0.41 (-0.80; -0.02) | 0.08 (-0.56; 0.71) |
| Power's ratio | -3.05 (-4.25; -1.82)* | 0.23 (-0.68; 1.13) | 0.51 (-0.92; 1.92) |
| D2L | -2.02 (-3.02; -0.98)* | -0.37 (-1.28; 0.55) | 0.16 (-1.25; 1.56) |
| Foramen magnum | 1.82 (0.81; 2.80)* | 0.51 (-0.42; 1.42) | -0.57 (-1.97; 0.87) |
| CD:FM | -2.17 (-3.20; -1.10)* | -0.08 (-0.98; 0.82) | 0.34 (-1.08; 1.74) |
| BDI [mm] | -1.27 (-2.17; -0.34)* | 0.42 (-0.50; 1.33) | -0.19 (-1.59; 1.22) |

| **Phenotyping**<br>(>16.5 years old) | Controls vs. Hurler | Controls vs.<br>Hurler-Scheie | Controls vs. MPS VI | Hurler vs.<br>Hurler-Scheie | Hurler vs. MPS VI |
| --- | --- | --- | --- | --- | --- |
| Cross-sectional area | 0.57 (0.16; 0.98)* | 3.31 (2.55; 4.05)* | 1.51 (0.98; 2.03)* | 2.06 (1.32; 2.79)* | 0.73 (0.18; 1.28)* |
| AP diameter | 0.51 (0.10; 0.91)* | 1.41 (0.80; 2.01)* | 1.25 (0.73; 1.76)* | 0.85 (0.21; 1.48)* | 0.70 (0.15; 1.24)* |
| RL diameter | 0.35 (-0.05; 0.75) | 3.36 (2.60; 4.10)* | 0.87 (0.37; 1.37)* | 2.74 (1.91; 3.55)* | 0.49 (-0.05; 1.03) |
| Power's ratio | -2.60 (-3.90; -1.26)* | -1.43 (-2.73; -0.08) | -3.87 (-5.79; -1.90)* | 1.29 (-0.14; 2.66) | -1.36 (-2.73; 0.09) |
| D2L | -1.08 (-2.08; -0.04) | -4.80 (-7.01; -2.53)* | -4.21 (-6.23; -2.13)* | -0.64 (-1.90; 0.65) | -1.58 (-3.02; -0.07) |

**Table 3 Effect sizes (Cohen's *d*) for cervical spinal cord quantitative MRI parameters across myelopathy severity categories and MPS genotypes**
| **Phenotyping**<br>(7.5-15.5 years old) | Controls vs. Hurler | Controls vs. Scheie | Hurler vs. Scheie |
| --- | --- | --- | --- |
| Fractional anisotropy | 1.37 (0.88; 1.85)* | -0.28 (-1.07; 0.51) | -1.33 (-2.17; -0.47)* |
| Radial diffusivity | -0.99 (-1.45; -0.52)* | 0.96 (0.14; 1.76)* | 1.35 (0.50; 2.20)* |
| Mean diffusivity | -0.38 (-0.82; 0.06) | 1.51 (0.67; 2.35)* | 1.22 (0.37; 2.05)* |
| Foramen magnum | 2.99 (1.22; 4.70)* | -1.23 (-3.11; 0.75) | -6.60 (-11.29; -1.90)* |
| CD:FM | -2.56 (-4.11; -0.93)* | 0.79 (-1.09; 2.59) | 3.79 (0.69; 6.79)* |

FA and AD were lower, and RD was higher, in MPS patients with stenosis compared with HC (**Fig. 3a**). RD was higher in MPS patients with SC compression than in those with stenosis (**Fig. 3a**). However, DTI did not show differences between MPS patients with myelopathy and those with compression (**Fig. 3a**). Effect sizes for DTI’s sensitivity to myelopathy severity are summarized in the first panel of **Table 3**. Overall, the morphologic and microstructural results objectively support our second hypothesis that CSC structure is sensitive to the severity of compression and myelopathy in MPS I and VI. Neither cervical neuroimaging measurement correlated with the PSS (Ahmed et al. 2016a), suggesting that the PSS is not sensitive to CSC pathology in MPS I and VI.

C4-C7 FA was positively correlated with body height in the recruited patient cohort, whereas C4-C7 RD and MD were negatively correlated (**Fig. 3b**). All correlations remained significant or showed a trend when accounting for age at scan as a confounding factor (**Fig. 3b**). Correlations between CSC DTI metrics and body height were not observed in the cohort of matched HC (**Supplementary Figure S9**) or in a concurrent large cohort of healthy young adults (Labounek et al. 2025), suggesting an MPS disease-related interaction. Overall, we may see weaker DTI-based CSC inner integrity in shorter patients (**Fig. 3b**). Cervical level-specific correlations are shown in **Supplementary Figure S10**. The foramen magnum dimension was also correlated with body height in MPS patients (r=0.54; p=0.013; p_A|Age_=0.031).

In the Hurler syndrome (n=13) and MPS VI (n=2) subcohorts treated with HSCT, the data showed a causal relationship between the age at HSCT and upper CSC morphology (**Fig. 3c**). Specifically, C1-C2 d_AP_ and C4 d_RL_ were lower in patients treated in later infancy or toddlerhood (**Fig. 3c**). The type of stem cell transplant (i.e., cord blood or bone marrow) and the type of donor (i.e., unrelated match or sibling) did not appear to affect these observed causal interactions (**Fig. 3c**). However, our sample size is too small to draw definitive conclusions. All causal relationships remained significant or showed a trend when accounting for age at scan as a confounding factor (**Fig. 3c**). Age at transplant and age at study scan acquisition were not correlated (r=-0.27; p=0.396).

CSC MRI in MPS can provide neuroimaging phenotypes, as shown by comparisons with age-matched patients with Hurler syndrome: (i) C1-C7 CSA, d_AP_ and d_RL_ differed in Hurler-Scheie syndrome (**Fig. 3d**); (ii) C1-C7 CSA and d_AP_ differed in MPS VI (**Fig. 3d**); and (iii) C1-C7 FA, RD and MD differed in Scheie syndrome (**Fig. 3e**). Moreover, most of these CSC phenotype measurements differed from those of HC (**Fig. 3d,e**). Cervical level-specific differences are shown in **Supplementary Figures S11-S12**. Dens-to-laminar interval (D2L) differed in MPS VI patients compared with those with Hurler syndrome **(Supplementary Figure S11**). Foramen magnum diameter and CD:FM differed in patients with Scheie syndrome when compared to Hurler syndrome (**Supplementary Figure S12**). Effect sizes for the phenotyping results are summarized in the second and third panels of **Table 3**. A larger MPS sample size may demonstrate the ability of Power’s ratio and D2L to differentiate among Hurler, Hurler-Scheie, and MPS VI genotypes (**Table 3**). We objectively confirm our third hypothesis that quantitative neuroimaging of CSC structure provides phenotypes sensitive to MPS genotypes.

## Discussion

We identified high-resolution structural neuroimaging in CSC as a potential source of biomarkers (Strimbu and Tavel 2010; Feng and Pepe 2020) for MPS I and VI because the investigated measurements: (i) differed from those in HC; (ii) were sensitive to the severity of degenerative compression and myelopathy; (iii) were sensitive to growth deficits in MPS patients; (iv) may be associated with HSCT responses related to treatment timing; and (v) were specific to MPS genotypes. However, larger cohorts and further validation within prospective longitudinal studies are needed.

Primary causes of cervical stenosis in MPS are characterized by two distinct mechanisms: Developmental skeletal dysplasia involves hypoplasia or aplasia of the odontoid process (dens axis), causing C0-2 hypermobility and atlantoaxial subluxation, which is especially severe in MPS IV and VI, followed by MPS I. Acquired soft-tissue storage affects the periodontoid tissue, ligamentum flavum, posterior longitudinal ligament, and dura mater (White and Harmatz 2010; Alden et al. 2017; Nestrasil and Vedolin 2017). In this work, we empirically measured cervical spinal canal narrowing, evaluated CSC compression and myelopathy, and assessed their effects on morphological and microstructural CSC measures using MRI. The main goal is to understand CSC pathology, predict potentially damaging changes, and prevent irreversible neurological deficits through noninvasive MRI surveillance of the CSC.

About 1%-2% of older adults (age 57±11 years) suffer from DCM (Kovalova et al. 2016; Badhiwala et al. 2020; Smith et al. 2021). In our MPS cohort, we detected myelopathy in 10% of cases at age 25 years. When we restricted our cohort to Hurler-Scheie syndrome, we estimated a myelopathy prevalence of 67%. This indicates that myelopathy is quite prevalent in MPS patients and begins in early life. Based on our work, CSC morphology is a candidate biomarker (Strimbu and Tavel 2010; Feng and Pepe 2020) with high sensitivity to the severity of SC compression and myelopathy, response to HSCT timing, and MPS genotype. The association between early HSCT and MRI findings or neurocognition aligns with our group’s previous findings in MPS I Hurler patients (Shapiro et al. 2015; Eisengart et al. 2017; Kovac et al. 2022; Svatkova et al. 2025).

Brain DTI studies have shown microstructural deficits in the corpus callosum in MPS I and VII (Shapiro et al. 2012; Kumar et al. 2014; Provenzale et al. 2015; Choi et al. 2015; Nestrasil et al. 2017; Nestrasil and Vedolin 2017; Svatkova et al. 2025); however, DTI of the spinal cord has not been explored in MPS yet. Therefore, we propose pioneering quantitative neuroimaging of microstructural myelopathy in MPS. We confirmed that DTI is a candidate biomarker for early myelopathy, as shown in DCM (Martin et al. 2017, 2018; David et al. 2019; Labounek et al. 2020; Valošek et al. 2021), traumatic SC injury (David et al. 2019), adrenoleukodystrophy (Huffnagel et al. 2019), amyotrophic lateral sclerosis (Pisharady et al. 2020), and Friedreich’s ataxia (Joers et al. 2022; Rezende et al. 2023). CSC DTI was sensitive to degenerative compression severity, growth deficits, and the Scheie syndrome genotype.

Neuroimaging data from young, healthy adults showed a moderate correlation between CSC CSA and body height and a potentially weak correlation between MD and body weight (Labounek et al. 2025). When we included body height as a confounding factor, some of our CSC structural measurements lost statistical significance. However, given the short stature of MPS patients, its proportionality to disease severity, and the correlation between microstructural measurements and body height, we conclude that the CSC structural measurements are more likely to reflect MPS mutations and related degenerative processes affecting both growth and the central nervous system.

Our study has several limitations. The limited sample size and relatively broad age range reflect the rarity of MPS I and VI. Although we demonstrate reasonable reproducibility of our MRI measurements in longitudinal data from several recruited patients, the overall study design is cross-sectional. Further prospective and longitudinal testing within clinical trials is needed to develop and validate subject-specific CSC neuroimaging biomarkers of SC health in MPS. In addition to recruiting more participants, future studies should enroll patients with other disease mutations, such as MPS II, III, IV, and VII. The reported PSS (Ahmed et al. 2016a, b) is not primarily designed or optimized to assess myelopathy severity. Therefore, further studies should record scores proportional to myelopathy severity, such as the modified Japanese Orthopedic Association score (Yonenobu et al. 1986; Benzel et al. 1991) or the neck disability index (Vernon and Mior 1991). The dMRI protocol may need improvement to increase sensitivity to myelopathy relative to measurements in degenerative compression. The currently used DTI-RESOLVE protocol (Labounek et al. 2020) was shown to provide lower sensitivity to microstructural myelopathy than the concurrent HARDI-ZOOMit protocol (Labounek et al. 2020; Valošek et al. 2021). The *spine-generic* DTI (Cohen-Adad et al. 2021) may be another concurrent dMRI protocol that improves sensitivity to manifested myelopathy. We show pilot sensitivity to varying MPS I and VI genotypes. However, each genotype underwent different treatment procedures. Therefore, there is a risk that our pilot results may be biased by different treatment effects. Studies of non-traumatic and traumatic SC injury show that myelopathy progresses rostrally above the focal lesion level and into supraspinal (brain) structures (Chen et al. 2017; Valošek et al. 2021; David et al. 2021; Yu et al. 2022; He et al. 2022; Wang et al. 2023; Schading et al. 2023). This remote degeneration is characterized by axonal loss, demyelination, and gray matter atrophy across multiple neuroanatomical levels. However, brain neuroimaging data were not utilized or associated with the CSC measurements in the current study. Additionally, the axial T_2_-weighted scans lacked sufficient contrast to delineate CSC gray matter. Transitioning to axial multi-echo MRI sequences (Labounek et al. 2020; Cohen-Adad et al. 2021, 2022) is therefore essential to resolve internal CSC morphology in greater anatomical detail.

## Data Availability

All data produced in the present study are available upon reasonable email request to Dr. Nestrasil.
In-house implemented computer code that provides automated image and statistical analyses is available at: https://github.com/umn-milab/mps-sc-resolve

## Artificial Intelligence Disclosure

During the preparation of this work, the authors used Google Gemini (v 3.1 Pro) exclusively to edit and polish the grammar and improve the manuscript’s readability. After using this tool/service, the authors reviewed and edited the content as needed and take full responsibility for the content of the publication.

## Computer Code Availability

In-house implemented computer code that provides automated image and statistical analyses is available at: https://github.com/umn-milab/mps-sc-resolve

## Data Availability

The data that support the findings of this study are available from Dr. Nestrašil upon reasonable email request.

## Author Contributions

IN contributed to study design, recruitment, data acquisition, curation and analysis, neurological assessment, and manuscript drafting and revision. CL contributed to study design, MRI protocol design, data analysis, and manuscript revision. IK contributed to manual morphological measurements and manuscript revision. BAM contributed to MRI protocol design, data curation, and manuscript revision. AA contributed to MPS severity assessment, mutation analysis, retrospective analysis of medical history, and manuscript revision. MTB contributed to recruitment, data acquisition and analysis, and manuscript revision. IL contributed to data analysis and manuscript revision. TC contributed to statistical analysis design and manuscript revision. AS contributed to study design, radiological reading of MRI data, and manuscript revision. CW contributed to mutation analysis and manuscript revision. RL contributed to study design, data acquisition, curation and analysis, figure design, manuscript drafting and revision, and overall team oversight.

## Acknowledgements

The Center for Magnetic Resonance Research (CMRR), Department of Radiology; and the Center for Neurobehavioral Development (CNBD) at the Masonic Institute for the Developing Brain (MIDB), Department of Pediatrics; all are part of the University of Minnesota and all provided lab space and computational support for this research project. Data were acquired at the CMRR under the support of the Lysosomal Disease Network.

Research reported in this publication was supported by the National Institute of Neurological Disorders and Stroke of the National Institutes of Health (NIH) under award number [R03NS139000], the Million Dollar Bike Ride from the University of Pennsylvania (MDBR-17-123-MPS), and the Rare Diseases Clinical Research Network, Lysosomal Disease Network, NIH U54NS065768, and the resources of the CMRR (supported by NIBIB P41 EB027061, P30 NS076408, and 1S10OD017974-01). The content is solely the responsibility of the authors and does not necessarily represent the official views of the NIH.

