## Supplementary Figures for "Structural damage of the cervical spinal cord within and beyond the maximal compression area in mucopolysaccharidosis types I and VI"

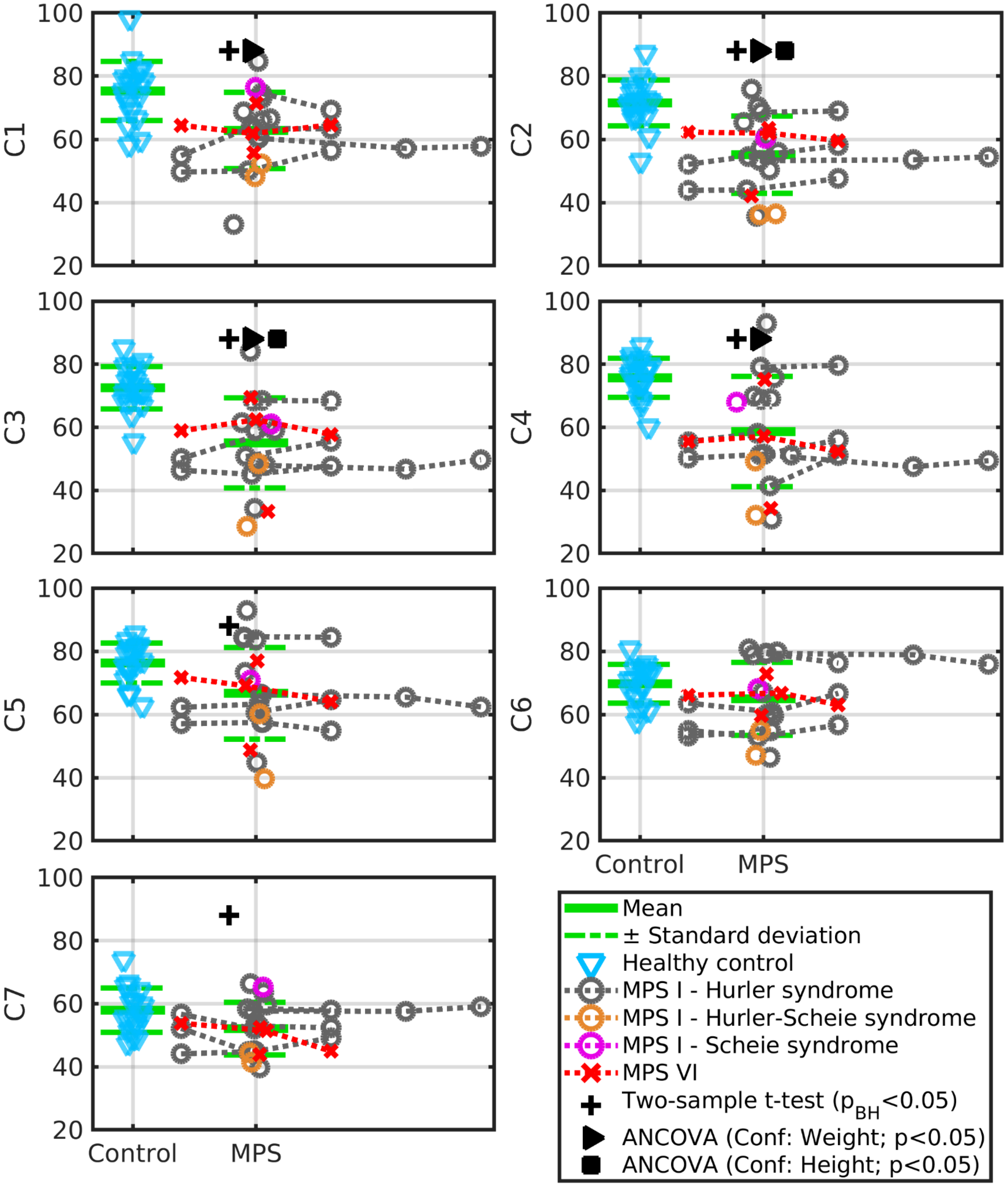


**Figure S1: Cross-sectional area of the cervical spinal cord at the C1-C7 levels in case-matched healthy controls and MPS I and VI patients.** Cross-sectional area is reported in $mm^2$. Healthy controls were sex- and age-matched to MPS patients at the visit, and group averages and standard deviation intervals are shown. Some MPS patients have longitudinal data acquired 1 year before the matched visit (i.e., measurements left of the matched visit), and some have longitudinal data acquired 1, 2, and/or 3 years after the matched visit (i.e., measurements right of the matched visit). Cross markers above the error bars indicate measurements in which two-sample t-tests with Benjamini-Hochberg adjusted p-values ($p_{BH}<0.05$) identified significant between-group differences. Triangle and square markers above the error bars indicate measurements in which post hoc analyses of covariance (ANCOVAs) using body weight or body height as a confounding factor identified significant differences between groups ($p<0.05$).

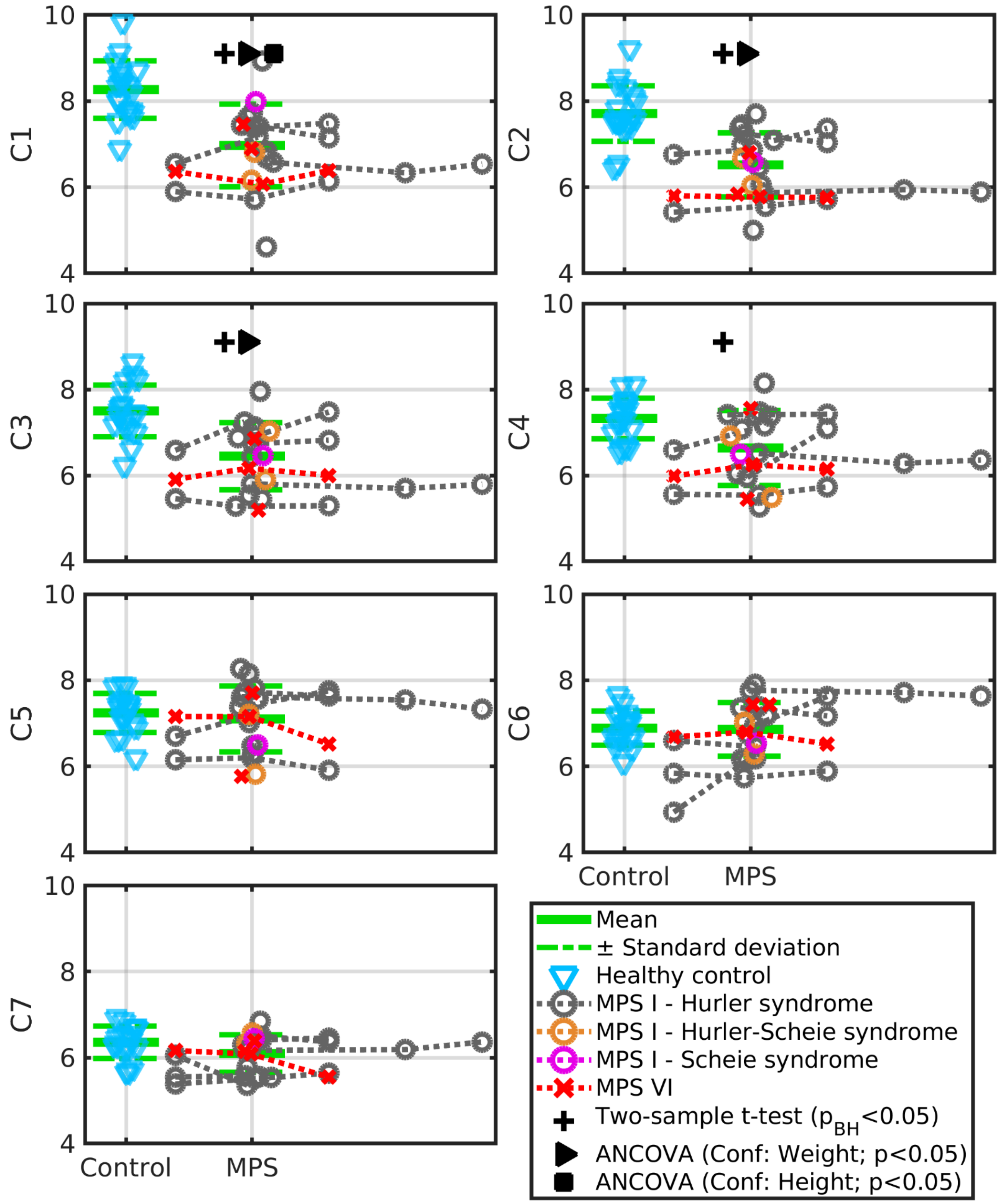


**Figure S2: Anterior-posterior diameter of the cervical spinal cord at the C1-C7 levels in case-matched healthy controls and MPS I and VI patients.** The diameter units are reported in mm. Healthy controls were sex- and age-matched to MPS patients at the visit, and group averages and standard deviation intervals are shown. Some MPS patients have longitudinal data acquired 1 year before the matched visit (i.e., measurements left of the matched visit), and some have longitudinal data acquired 1, 2, and/or 3 years after the matched visit (i.e., measurements right of the matched visit). Cross markers above the error bars indicate measurements in which two-sample t-tests with Benjamini-Hochberg adjusted p-values ($p_{BH}<0.05$) identified significant between-group differences. Triangle and square markers above the error bars indicate measurements in which post hoc analyses of covariance (ANCOVAs) using body weight or body height as a confounding factor identified significant differences between groups ($p<0.05$).

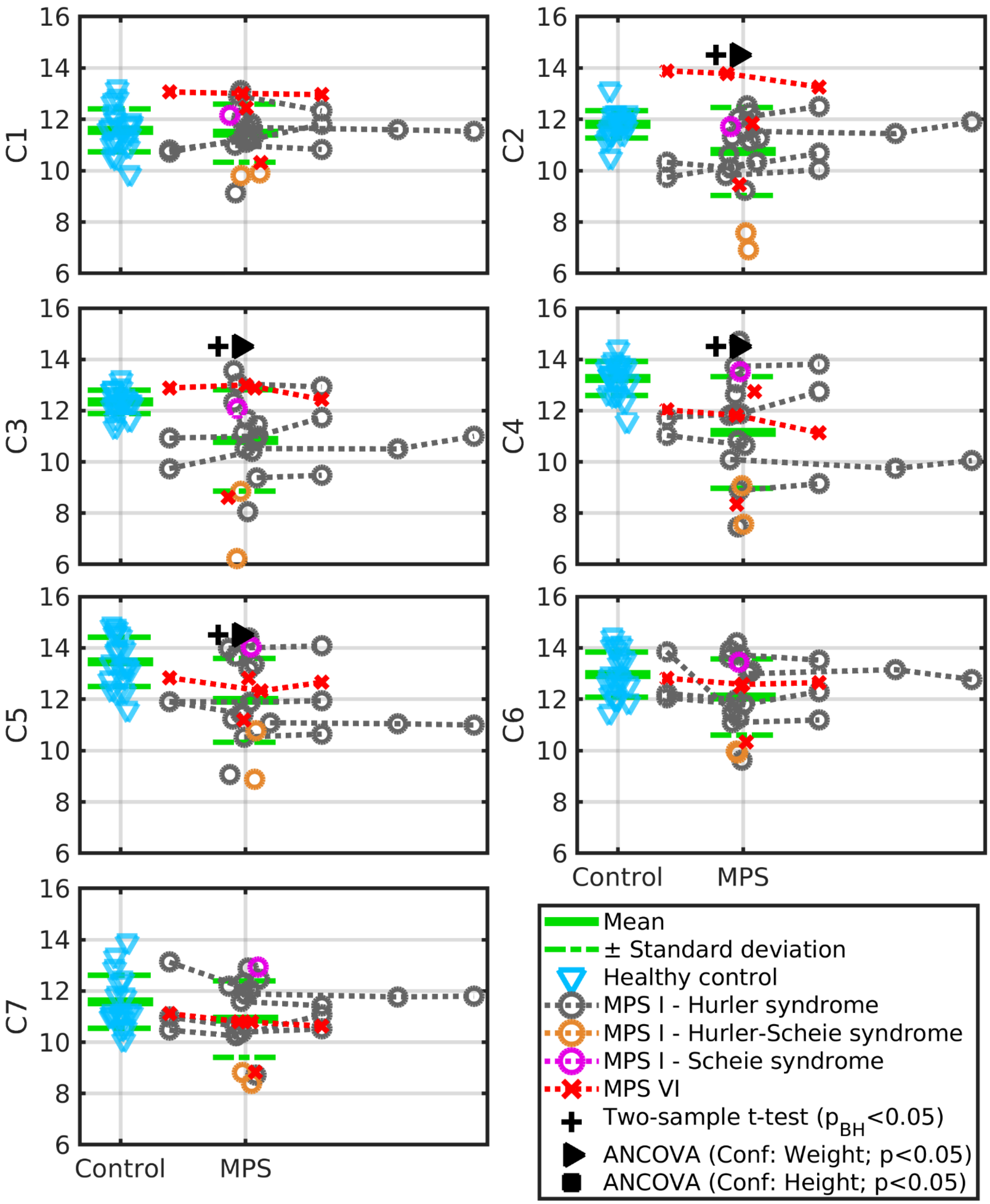


**Figure S3: Right-left diameter of the cervical spinal cord at the C1-C7 levels in case-matched healthy controls and MPS I and VI patients.** The diameter units are reported in mm. Healthy controls were sex- and age-matched to MPS patients at the visit, and group averages and standard deviation intervals are shown. Some MPS patients have longitudinal data acquired 1 year before the matched visit (i.e., measurements left of the matched visit), and some have longitudinal data acquired 1, 2, and/or 3 years after the matched visit (i.e., measurements right of the matched visit). Cross markers above the error bars indicate measurements in which two-sample t-tests with Benjamini-Hochberg adjusted p-values ($p_{BH}<0.05$) identified significant between-group differences. Triangle and square markers above the error bars indicate measurements in which post hoc analyses of covariance (ANCOVAs) using body weight or body height as a confounding factor identified significant differences between groups ($p<0.05$).

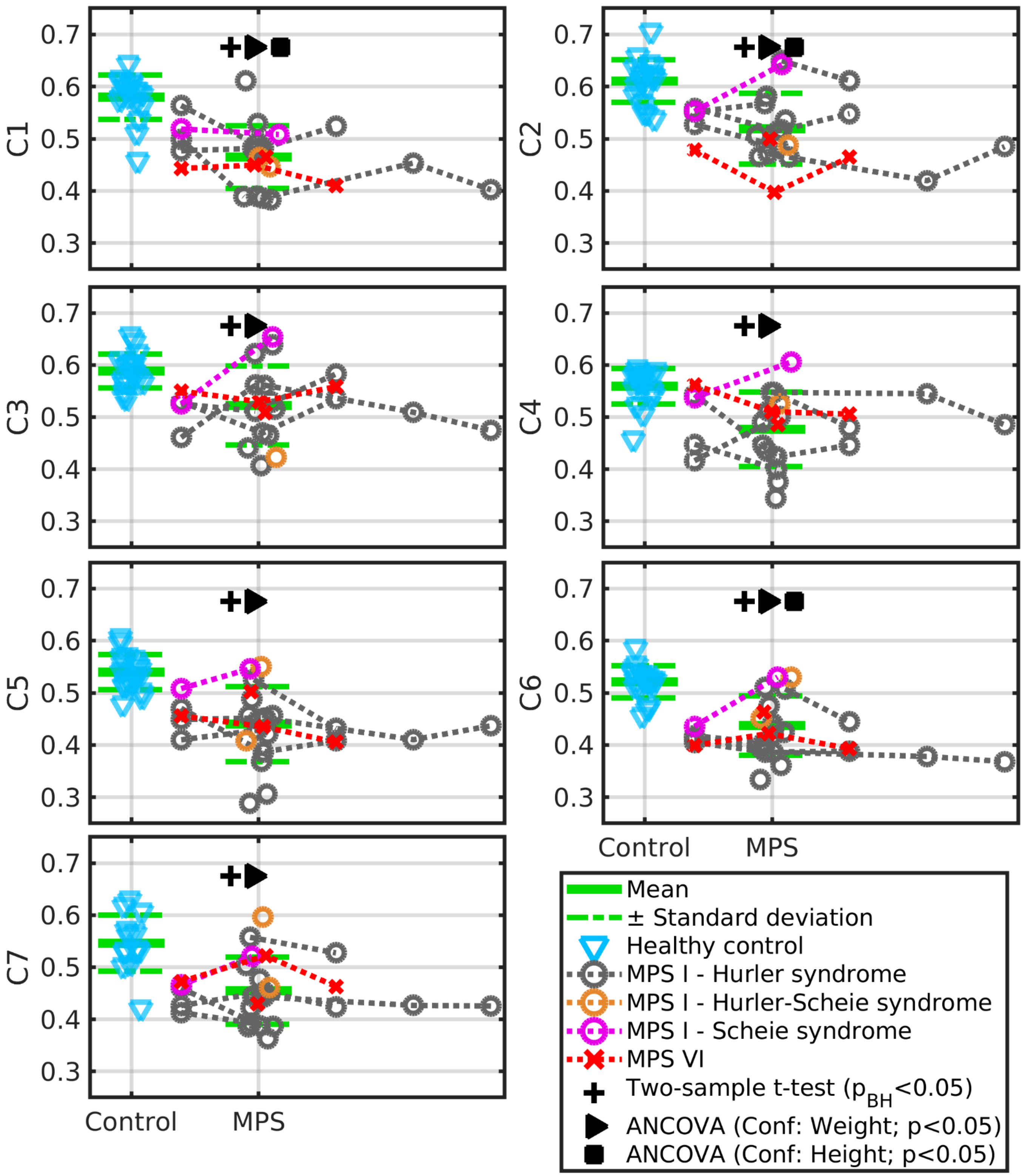


**Figure S4: Fractional anisotropy of the cervical spinal cord at the C1-C7 levels in case-matched healthy controls and MPS I and VI patients.** Healthy controls were sex- and age-matched to MPS patients at the visit, and group averages and standard deviation intervals are shown. Some MPS patients have longitudinal data acquired 1 year before the matched visit (i.e., measurements left of the matched visit), and some have longitudinal data acquired 1, 2, and/or 3 years after the matched visit (i.e., measurements right of the matched visit). Cross markers above the error bars indicate measurements in which two-sample t-tests with Benjamini-Hochberg adjusted p-values ($p_{BH}<0.05$) identified significant between-group differences. Triangle and square markers above the error bars indicate measurements in which post hoc analyses of covariance (ANCOVAs) using body weight or body height as a confounding factor identified significant differences between groups ($p<0.05$).

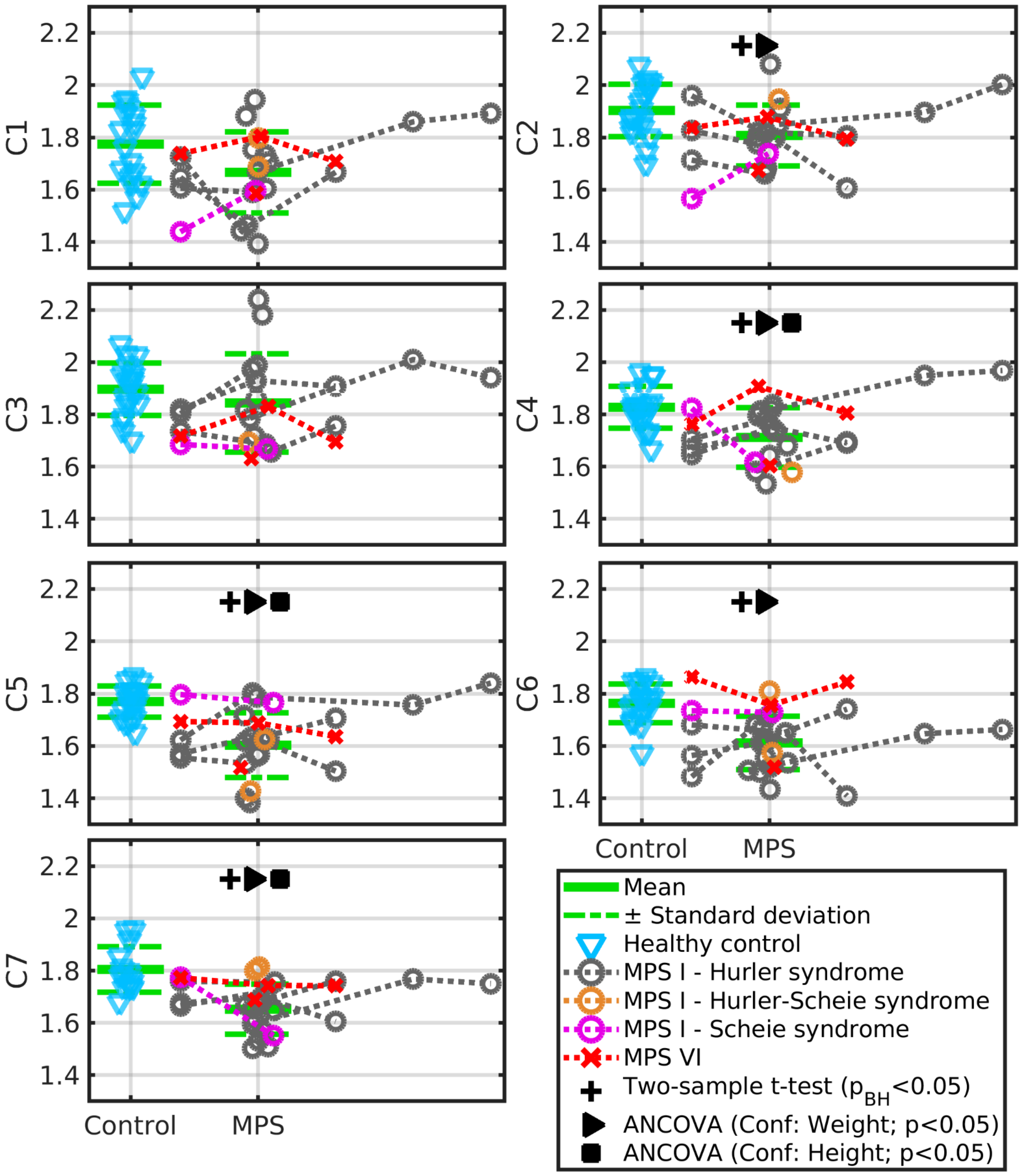


**Figure S5: Axial diffusivity of the cervical spinal cord at the C1-C7 levels in case-matched healthy controls and MPS I and VI patients.** The axial diffusivity units are reported in $*10^{-9}m^2/s$. Healthy controls were sex- and age-matched to MPS patients at the visit, and group averages and standard deviation intervals are shown. Some MPS patients have longitudinal data acquired 1 year before the matched visit (i.e., measurements left of the matched visit), and some have longitudinal data acquired 1, 2, and/or 3 years after the matched visit (i.e., measurements right of the matched visit). Cross markers above the error bars indicate measurements in which two-sample t-tests with Benjamini-Hochberg adjusted p-values ($p_{BH}<0.05$) identified significant between-group differences. Triangle and square markers above the error bars indicate measurements in which post hoc analyses of covariance (ANCOVAs) using body weight or body height as a confounding factor identified significant differences between groups ($p<0.05$).

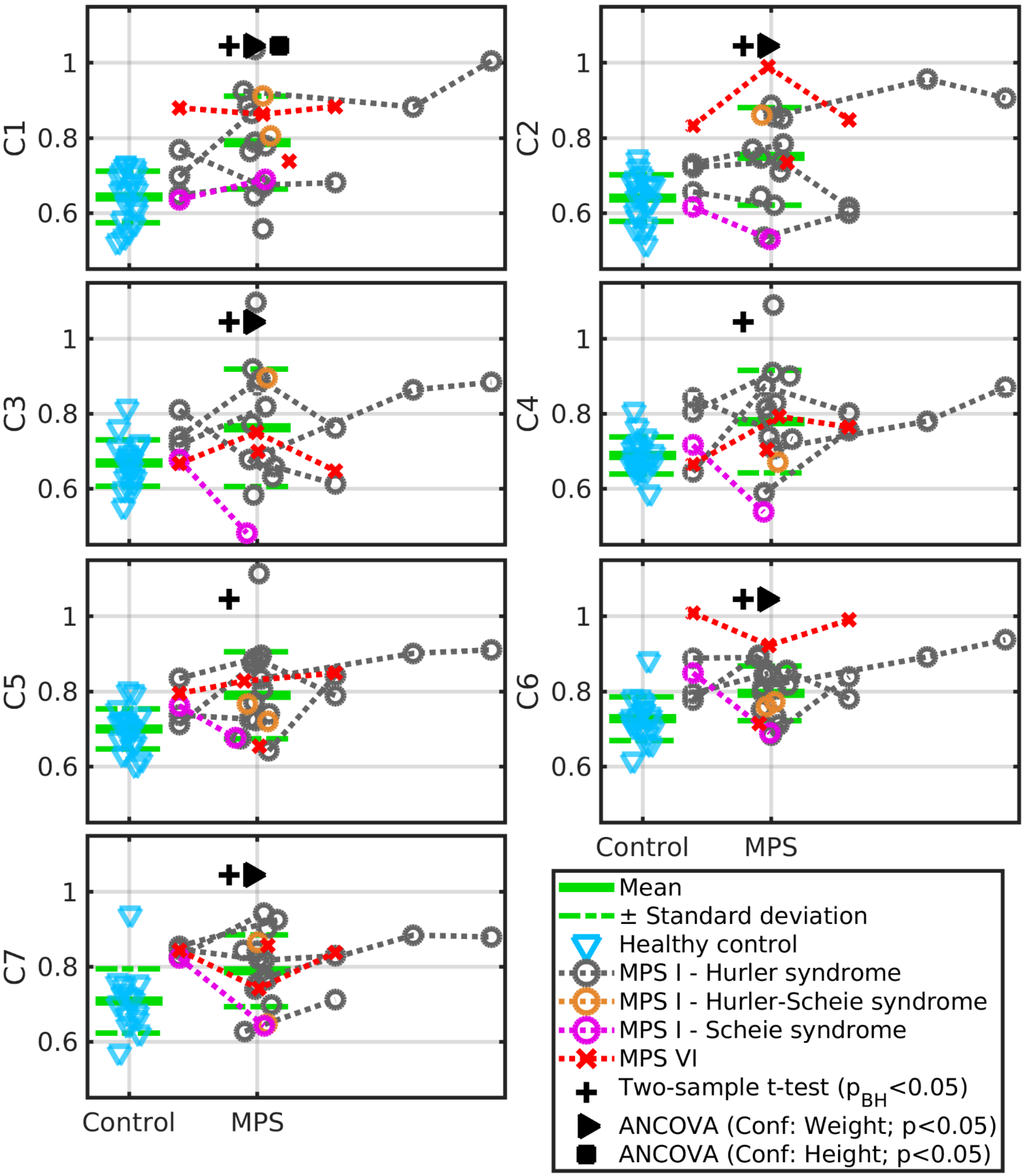


**Figure S6: Radial diffusivity of the cervical spinal cord at the C1-C7 levels in case-matched healthy controls and MPS I and VI patients.** The radial diffusivity units are reported in $*10^{-9}m^2/s$. Healthy controls were sex- and age-matched to MPS patients at the visit, and group averages and standard deviation intervals are shown. Some MPS patients have longitudinal data acquired 1 year before the matched visit (i.e., measurements left of the matched visit), and some have longitudinal data acquired 1, 2, and/or 3 years after the matched visit (i.e., measurements right of the matched visit). Cross markers above the error bars indicate measurements in which two-sample t-tests with Benjamini-Hochberg adjusted p-values ($p_{BH}<0.05$) identified significant between-group differences. Triangle and square markers above the error bars indicate measurements in which post hoc analyses of covariance (ANCOVAs) using body weight or body height as a confounding factor identified significant differences between groups ($p<0.05$).

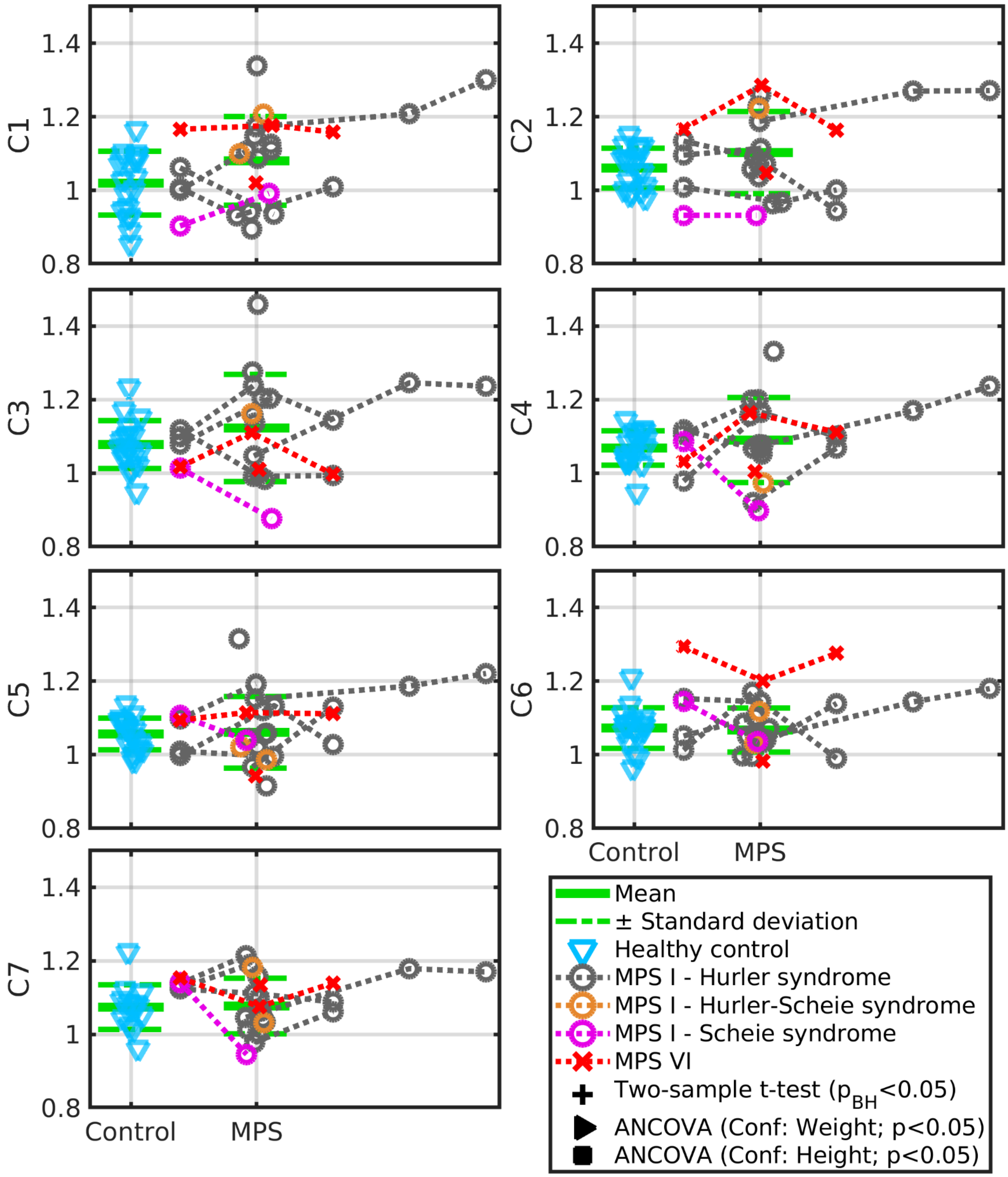


**Figure S7: Mean diffusivity of the cervical spinal cord at the C1-C7 levels in case-matched healthy controls and MPS I and VI patients.** The mean diffusivity units are reported in $*10^{-9} m^2/s$. Healthy controls were sex- and age-matched to MPS patients at the visit, and group averages and standard deviation intervals are shown. Some MPS patients have longitudinal data acquired 1 year before the matched visit (i.e., measurements left of the matched visit), and some have longitudinal data acquired 1, 2, and/or 3 years after the matched visit (i.e., measurements right of the matched visit). Cross markers above the error bars indicate measurements in which two-sample t-tests with Benjamini-Hochberg adjusted p-values ($p_{BH}<0.05$) identified significant between-group differences. Triangle and square markers above the error bars indicate measurements in which post hoc analyses of covariance (ANCOVAs) using body weight or body height as a confounding factor identified significant differences between groups ($p<0.05$).

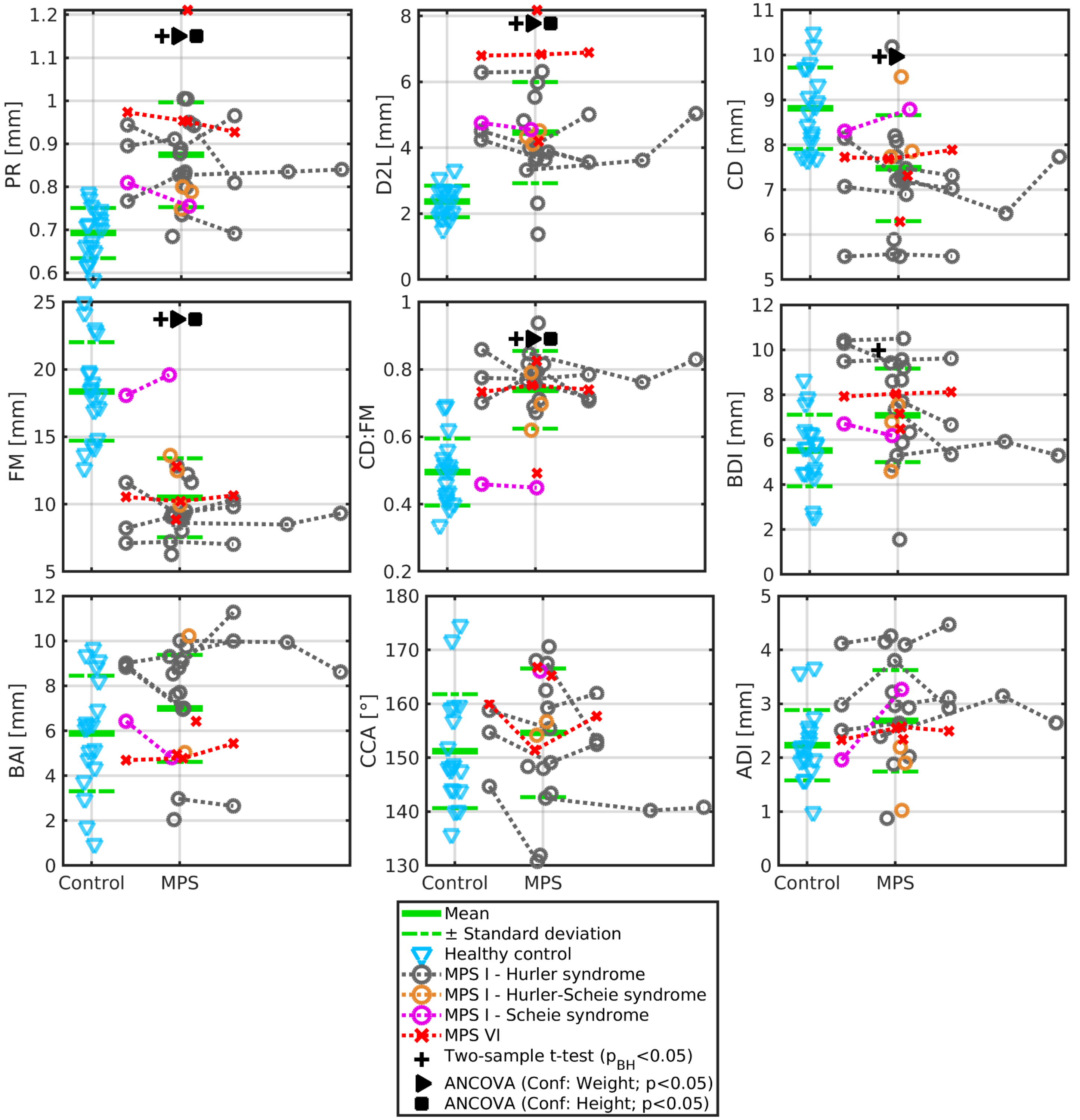


**Figure S8: Manual measurements in the occipito-cervical junction area in case-matched healthy controls and MPS I and VI patients.** Healthy controls were sex- and age-matched to MPS patients at the visit, and group averages and standard deviation intervals are shown. Some MPS patients have longitudinal data acquired 1 year before the matched visit (i.e., measurements left of the matched visit), and some have longitudinal data acquired 1, 2, and/or 3 years after the matched visit (i.e., measurements right of the matched visit). Cross markers above the error bars indicate measurements in which two-sample t-tests with Benjamini-Hochberg adjusted p-values ($p_{BH}<0.05$) identified significant between-group differences. Triangle and square markers above the error bars indicate measurements in which post hoc analyses of covariance (ANCOVAs) using body weight or body height as a confounding factor identified significant differences between groups ($p<0.05$).

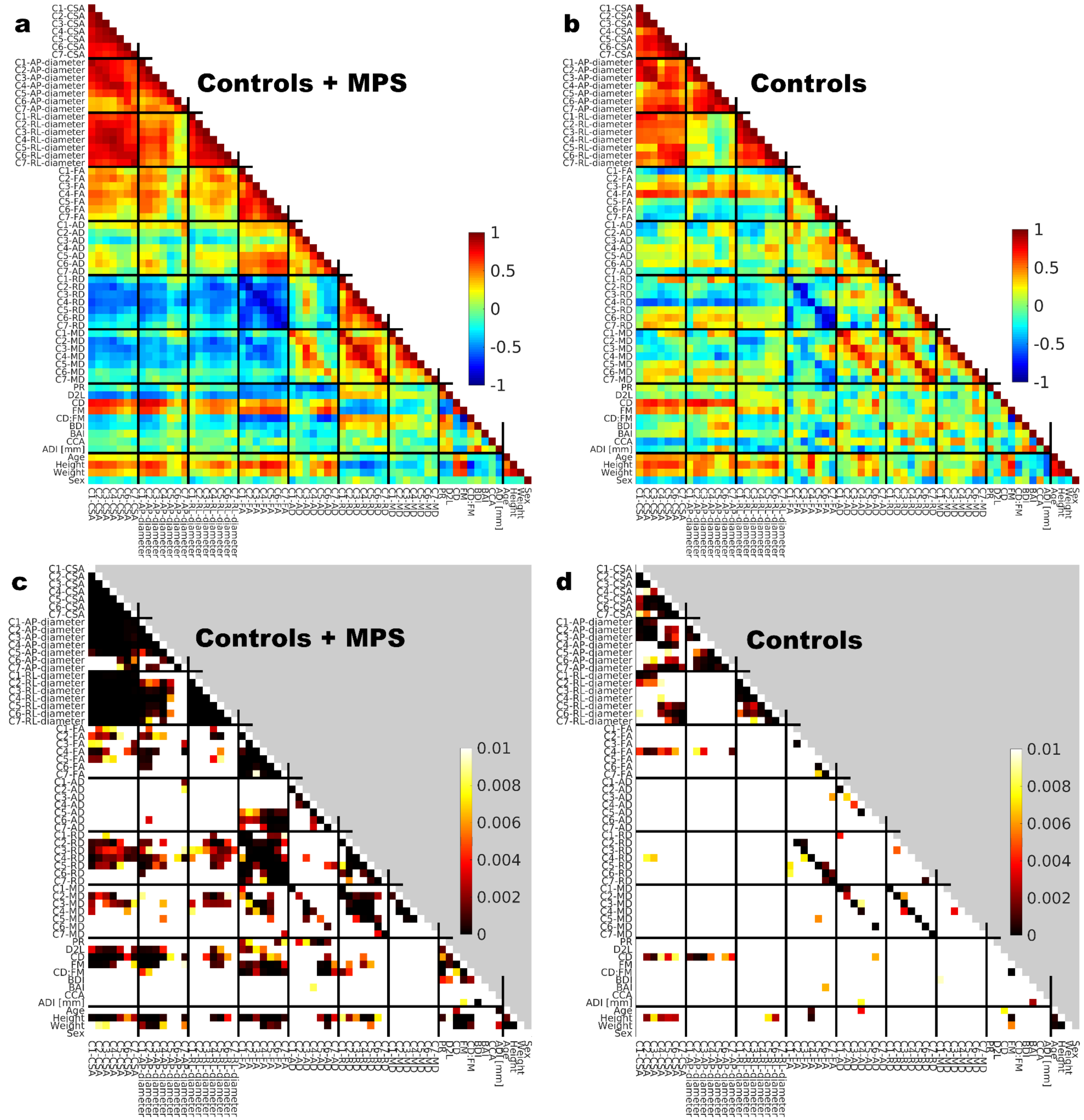


**Figure S9: Cross-correlation matrices of investigated variables for case-matched healthy controls and MPS I and VI patients.** **a**, Matrix of Pearson correlation coefficients for healthy controls and MPS patients together. **b**, Matrix of Pearson correlation coefficients for healthy controls only. **c**, Matrix of p-values of Pearson correlation coefficients for healthy controls and MPS patients together. **d**, Matrix of p-values of Pearson correlation coefficients for healthy controls only. Structure of the correlation matrices differs when assembled for controls+patients or controls only.

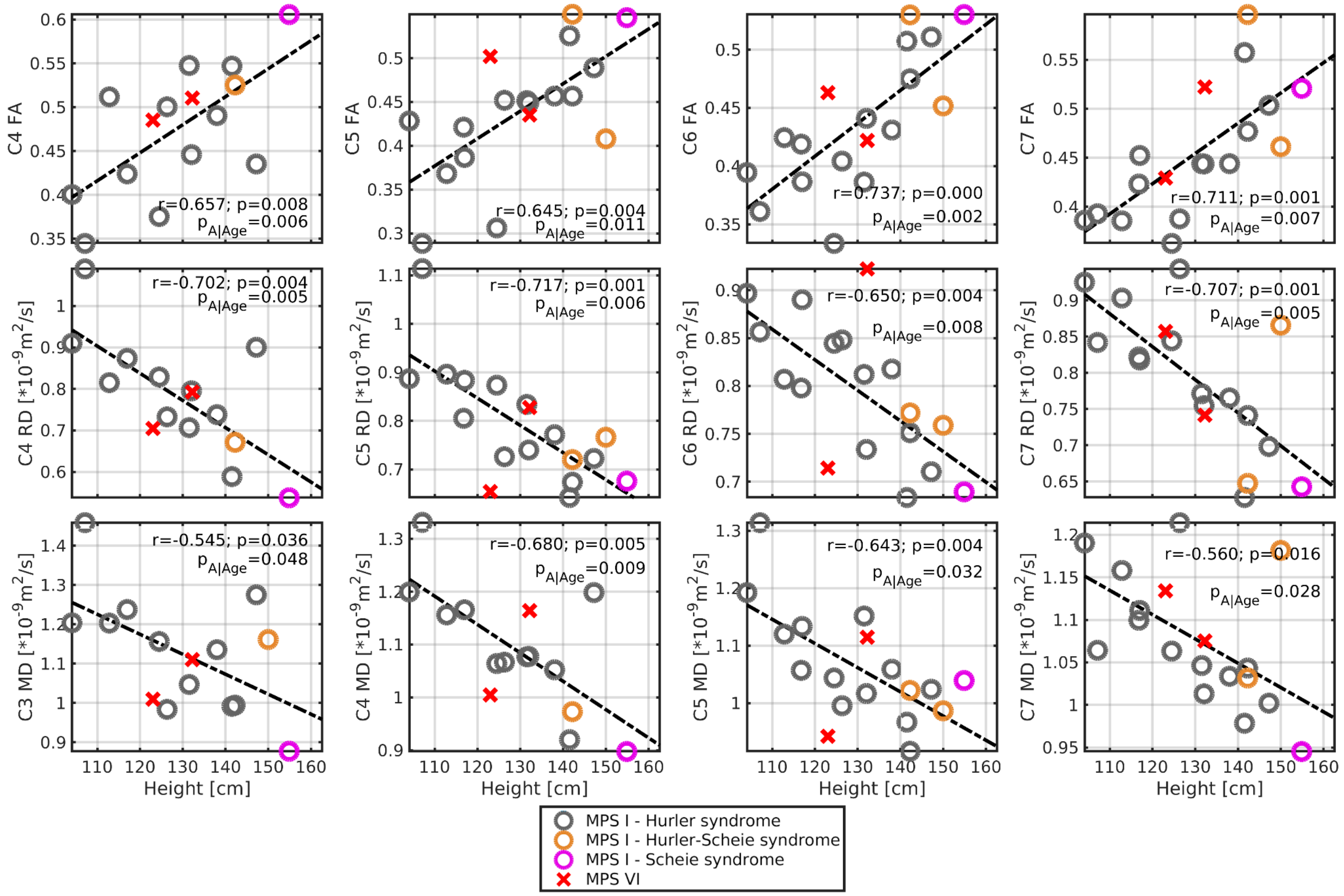


**Figure S10: Pearson correlation coefficients (*r*) with corresponding p-value (*p*) between mid and lower CSC DTI and body height in MPS I and VI patients.** The $p_{A|Age}$ demonstrates the p-value of the relationship when age at scan is utilized as the confounding factor in analysis of covariance.

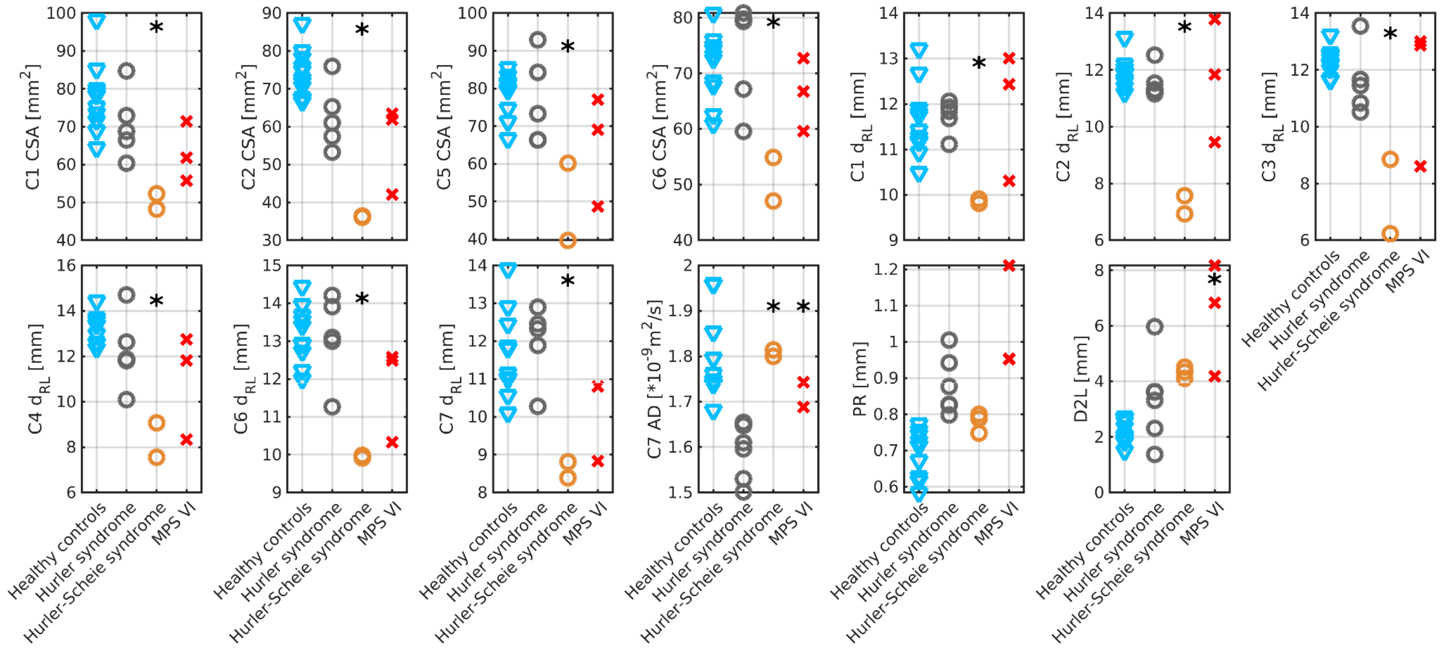


**Figure S11: Genotype-specific neuroimaging measurements.** The categorical scatter plots show subsets of age-matched data samples (i.e., three 20-30 years old patients with Hurler-Scheie syndrome and three 20-30 years old MPS VI patients). The between-genotype differences were compared to Hurler syndrome data with two-sample t-tests ($p<0.05$) and marked with the star above the other genotypes that differed in morphological measurements.

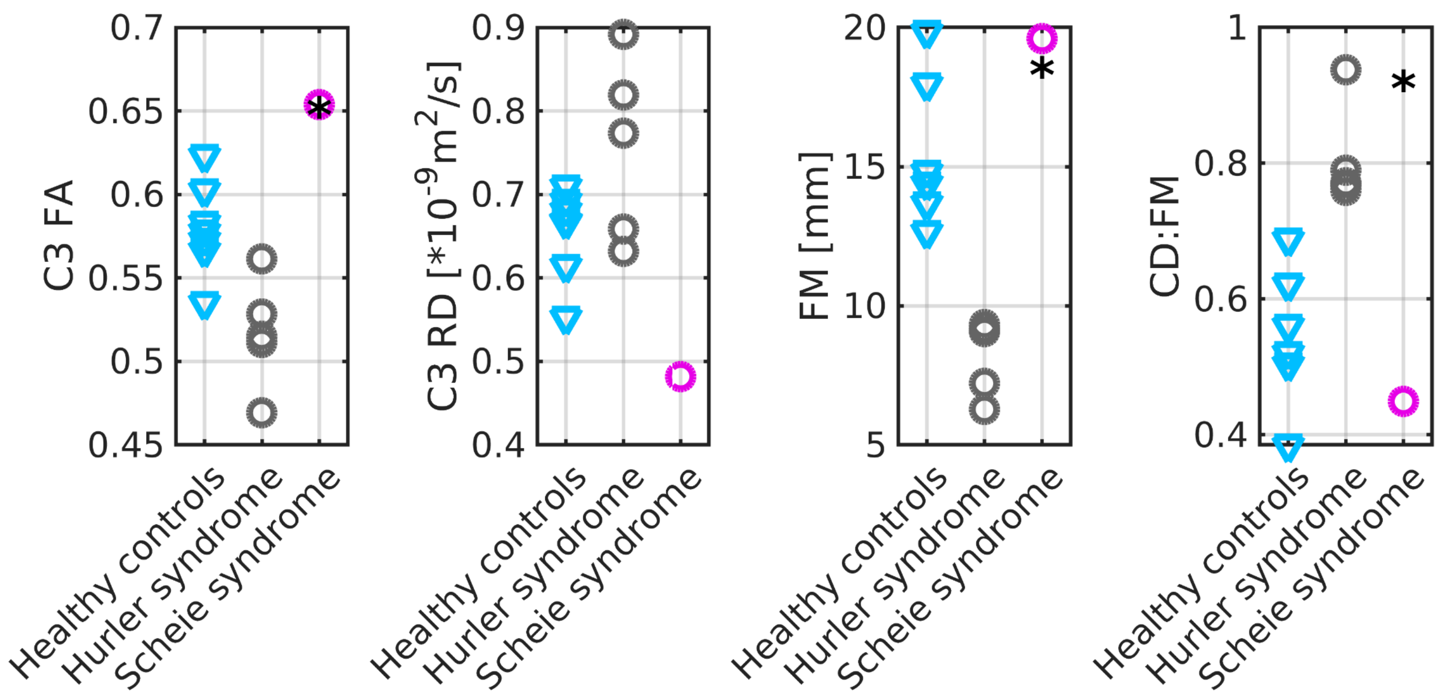


**Figure S12: Genotype-specific neuroimaging measurements.** The categorical scatter plots show subsets of age-matched data samples. The between-genotype differences were compared to Hurler syndrome data with two-sample t-tests ($p<0.05$) and marked with the star above the other genotypes that differed in morphological or DTI measurements.
